# Modeling early-onset cancer kinetics to study changes in underlying risk, detection, and impact of population screening

**DOI:** 10.1101/2024.12.05.24318584

**Authors:** Navid Mohammad Mirzaei, Chin Hur, Mary Beth Terry, Piero Dalerba, Wan Yang

## Abstract

Recent studies have reported increases in early-onset cancer cases (diagnosed under age 50) and raised questions about whether the increase is related to earlier diagnosis from non-specific medical tests as reflected by decreasing tumor-size-at-diagnosis (apparent effects) or actual increases in underlying cancer risk (true effects), or both. The classic Multi-Stage Clonal Expansion (MSCE) model assumes cancer detection at the first malignant cell’s emergence, although later modifications have included lag-times or stochasticity in detection to represent the delay in tumor detection. Here, we introduce an approach to explicitly incorporate tumor-size-at-diagnosis in the MSCE framework accounting for improvements in cancer detection over time to distinguish between apparent and true increases in early-onset cancer incidence. We demonstrate that our model is structurally identifiable and provides better parameter estimation than the classic model. Applying this model to colorectal, breast, and thyroid cancers, we examine changes in cancer risk while accounting for detection improvements over time in three representative birth cohorts (1950-1954, 1965-1969, and 1980-1984). Our analyses suggest accelerated carcinogenic events and shorter mean sojourn times (the average time from the first malignant cell emergence to cancer detection) in more recent cohorts. We further use this model to examine the screening impact on the incidence of breast and colorectal cancers, both having established screening protocols. Our results align with well-documented differences in screening effects between these cancers. These findings underscore the importance of incorporating tumor-size-at-diagnosis in cancer modeling and support true increases in early-onset cancer risk in recent years for breast, colorectal, and thyroid cancer.

**Significance:** This study models recent increases in early-onset cancers, accounting for both true factors contributing to cancer risk and those caused by improved detection. We show that while advancement in detection has led to earlier detection, our model estimates shorter sojourn times and more aggressive carcinogenic events for recent cohorts, suggesting faster tumor progression. Further, a counterfactual analysis using this model reveals the known statistically significant reduction in colorectal cancer incidence (supporting a robust modeling approach), likely due to screening and timely removal of precancerous polyps. Overall, we introduce an enhanced model to detect subtle trends in cancer risk and demonstrate its ability to provide valuable insights into cancer progression and highlight areas for future refinement and application.

## Introduction

Cancer is the second most common cause of death in the world and is estimated to be responsible for about 2 million new cases and more than 600 thousand deaths in 2024, according to the American Cancer Society Statistics [1]. Moreover, globally, early-onset (less than 50 years of age) cancer incidence has increased by 79.1% with a 27.7% increase in death between 1990 and 2019 [2]. Many studies have aimed to understand the reason for such recent increases in early-onset incidence, with dietary factors, tobacco use, and alcohol consumption recognized as the most common modifiable risk factors [3]. Moreover, screening policies have been modified in response. For example, the United States Preventive Services Task Force (USPSTF) updated their guidelines in 2024, recommending that all women begin screening for breast cancer (BrC) at age 40, as opposed to ages 45 and 50 as recommended previously [4]. The USPSTF also recommended colorectal cancer (CRC) screening starting at age 45, lowered from 50, in 2021 [5]. Given that screening facilitates earlier diagnosis, such population-level screening could result in increases in incident cases. In addition, advancements in and increased use of medical imaging technologies such as CT scan [6] enable the detection of smaller tumors, even for cancer types with no population screening such as thyroid cancer (ThC) [7, 8], which could partly contribute to the apparent increases in incidence among young adults. Indeed, the tumor sizes recorded at diagnosis in general decreased over time since 1988, when the Surveillance, Epidemiology, and End Results (SEER) program started to report these data [9] (Figure 1(A)). Thus, the apparent increases need to be accounted for to gauge the true risk increases more accurately and to examine the underlying etiology of early-onset cancers.

**Figure 1.**
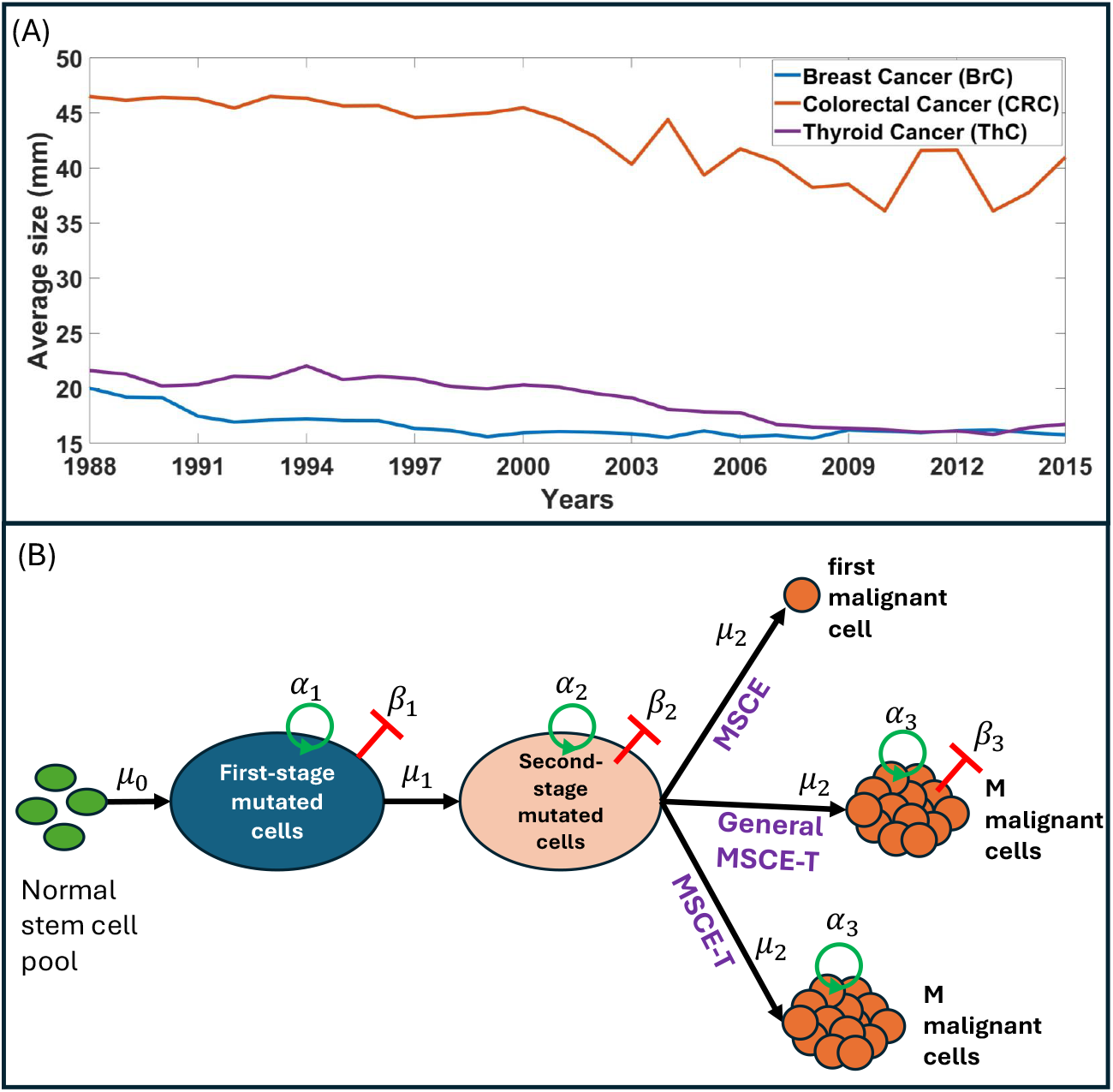
Motivation and schematic of the models. (A) The mean tumor-size-at-diagnosis for breast, colorectal, and thyroid cancer, extracted from the SEER database. (B) The carcinogenesis process from normal stem cells to malignant cells. The classic MSCE model records the cancer incidence at the time of the first malignancy occurrence. The General MSCE-T model considers birth (*α*_3_) and death (*β*_3_) rates for malignant cells and records the cancer incidence when the number of malignant cells reaches ≥ *N*. The main model (MSCE-T) considers a known proliferation rate (*α*_3_) for malignant cells and records the cancer incidence when the number of malignant cells reaches ≥ *N*.

Mathematical models have been applied to explore the underlying factors contributing to cancer incidence. A prime example of such application is Armitage and Doll’s pioneering work in 1954, which introduced the theory of multistage carcinogenesis and set the groundwork for similar cancer models [10]. This theory posits that cancer is the product of the accumulation of genetic mutations in normal cells. Building on this, in a series of papers, Moolgavkar et al. (1979, 1981) showed that a two-stage mutation model is insufficient to capture the intricacies of cancer incidence unless stage-wise clonal expansion of mutated cells is also considered [11, 12]. This led to the introduction of the Multistage Clonal Expansion (MSCE) model, which incorporates the proliferation of mutated cells at each stage in addition to the accumulation of genetic mutations. In recent years, various studies have modified and adopted the MSCE model to capture different details that contribute to cancer incidence. These include multiple studies examining the number of mutations (i.e., the number of stages in the MSCE model) needed to capture the age-specific incidence patterns for different cancer types [13, 14, 15, 16]; the development of different hazard functions to better capture the phases and transitions of tumor growth kinetics over the life course [17, 18]; additional model components to account for cancer detection [19, 20]; and the incorporation of population-or individual-level risk factors in the mutation rates [21, 22]. Another line of cancer kinetics modeling examines the evolutionary dynamics of cancer initiation and progression; for instance, Paterson et al. (2020) [23] and Li et al. (2023) [24] considered the heterogeneity of mutation orders to explore the genetic pathways that drive these dynamics.

Nonetheless, to date, MSCE models have yet to directly account for changes to cancer detection over time and the impact on observed cancer incidence, which is essential to understanding the reasons behind the recent early-onset cancer incidence increases as noted above. Here, we propose an approach to extend the MSCE models to capture the clonal expansion of malignancy beyond the first malignant cell (the endpoint in classic MSCE models) and account for changes in detection using data on tumor-size-at-diagnosis. To do so, the proposed model additionally represents the clonal expansion of malignant cells through the time of tumor diagnosis (per reported tumor-size-at-diagnosis), using a simple birth process in the MSCE framework. By adding this factor, our model encompasses both the apparent and true effects in the observed early-onset cancer incidence. Further, unlike previous works that numerically test the fitness of MSCE models with different numbers of stages [13, 14, 15, 16], we adopt a general modeling framework with three stages of mutation and clonal expansion based on findings from recent genetic analyses [25]. To capture the heterogeneity of cancers, we use incidence data to estimate the rate of progression during each stage and the relative contribution of each stage, such that the model can be applied to different cancer types.

To test the proposed approach, here we focus on three early-onset cancers with varying detection improvements and screening practices in the US: BrC, CRC, and ThC. These three cancers are among those with the most dramatic surges in early-onset incidence [3]. BrC and CRC are both recommended for population screening, but case detection prior to the recommended screening age is typically due to symptoms, despite improved diagnostics over time. In comparison, ThC is not recommended for population screening in the US. However, as noted above, the recent increased use of medical imaging and subsequent incidental detection of ThC has been cited as a reason for its rising incidence [7]. These differences allow model testing and comparison of the estimated apparent effects due to increased detection (more prominent for ThC compared to the other two cancer types) and true cancer risks, as well as examining the impact of population screening. We apply the proposed model to the three key early-onset cancers (BrC, CRC, and ThC) over multiple birth cohorts. Our results show that our model is fully structurally identifiable, has a better parameter estimation capability, and explains the increase in early-onset cancer better than the classic model. By accounting for changing detection, we are able to estimate the increases in tumor growth rates and the key stages of such increases in different birth cohorts for the three cancers. In addition, using the model estimates, we calculate the mean sojourn times (the average time from the occurrence of the first malignant cell to cancer detection) and conduct a counterfactual analysis to investigate the impact of population screening on BrC and CRC.

## The model: framework, identifiability and validation

### Model framework

It is a widely accepted theory that cancer arises from the accumulation of genetic mutations that turn normal stem cells into malignant ones [26]. Key mutations occur in genes that regulate cell proliferation and death [27], such as APC, TP53, and KRAS in colorectal cancer; TP53, PIK3CA, and GATA3 in breast cancer; and BRAF, CHEK2, and RET in thyroid cancer [28, 29, 30]. Research by Tomasetti et al. suggests that as few as three driver mutations may be sufficient for cancer to develop, based on studies of colorectal and lung cancers, which have the highest number of somatic mutations (i.e., can serve as an upper bound for other cancer types) [25]. Therefore, in this study, we consider three mutations before malignancy, but note the model provided here can be easily expanded to account for more mutations. We consider three models (see Table 1 for a summary). As shown in Figure 1 (B), the models assume normal stem cells mutate into First-stage Mutated Cells (FSMCs) at rate *µ*_0_, which may proliferate, die, or progress to Second-stage Mutated Cells (SSMCs) at rates *α*_1_, *β*_1_, and *µ*_1_, respectively; SSMCs similarly transition to malignancy at rates *α*_2_, *β*_2_, and *µ*_2_. The classic MSCE model [12] defines cancer incidence as the emergence of the first malignant cell. In contrast, the proposed MSCE-T (T for tumor) models define incidence when the malignant cells reach a threshold population (reflecting tumor size at diagnosis), enabling separation of apparent effects (caused by improved diagnosis over time) and true effects (caused by mutagenic factors). In addition to the parameters discussed above, the General MSCE-T model includes malignant cell proliferation (*α*_3_) and death (*β*_3_), while our main MSCE-T model represents net proliferation by keeping *α*_3_ and setting *β*_3_ = 0. We describe each model briefly below and provide the details in the Supplementary Materials.

**Table 1.** Models, model variables and parameters definition.

| Variable/Parameter/Model | Description |
| --- | --- |
| $x_1$ | Survival* given $N_0$ normal cells and no other cell types at time zero. |
| $x_2$ | Hazard rate** of cancer incidence, i.e., $-\frac{d}{dt}[\ln x_1]$ |
| $x_3$ | Survival given $N_0$ normal cells, one FSMC<sup>†</sup>, and no other cell types at time zero. |
| $x_4$ | Auxiliary variable defined as $\frac{d}{dt}[x_3]$ |
| $x_5$ | Survival given $N_0$ normal cells, one SSMC<sup>‡</sup>, and no other cell types at time zero. |
| $x_6$ | Auxiliary variable defined as $\frac{d}{dt}[x_5]$ |
| $N_0$ | $N_0 = 1.74 \times 10^{10}$ for breast [37],<br>$N_0 = 2 \times 10^8$ for colon and rectum [38],<br>and $N_0 = 6.5 \times 10^7$ for thyroid [38]. |
| $\mu_0$ | Mutation rate of normal/stem cells |
| $\mu_1$ | Mutation rate of FSMCs |
| $\mu_2$ | Mutation rate of SSMCs |
| $\alpha_1$ | Birth rate of FSMCs |
| $\alpha_2$ | Birth rate of SSMCs |
| $\alpha_3$ | Birth rate of malignant cells used in both MSCE-T models |
| $\beta_1$ | Death rate of FSMCs |
| $\beta_2$ | Death rate of SSMCs |
| $\beta_3$ | Death rate of malignant cells used only in the General MSCE-T model |
| $M_t$ | Number of malignant cells at diagnosis time $t$ |
| $\mu_1 \times (\alpha_1 - \beta_1)$ | Aggressiveness<sup>◊</sup> of FSMCs |
| $\mu_2 \times (\alpha_2 - \beta_2)$ | Aggressiveness of SSMCs |
| MSCE model | Equations (4)-(9) with $f(t) = 0$,<br>$[x_1(0), x_2(0), x_3(0), x_4(0), x_5(0), x_6(0)] = [1, 0, 1, 0, 1, -\mu_2]$,<br>cancer incidence := the occurrence of the first malignant cell,<br>No $\alpha_3$ and $\beta_3$. |
| General MSCE-T model | Equations (4)-(9) with $f(t)$ given by (10),<br>$[x_1(0), x_2(0), x_3(0), x_4(0), x_5(0), x_6(0)] = [1, 0, 1, 0, 1, 0]$,<br>cancer incidence := having $M_t$ or more malignant cells,<br>Both $\alpha_3$ and $\beta_3$ require estimation. |
| MSCE-T model | Equations (4)-(9) with $f(t)$ given by (11),<br>$[x_1(0), x_2(0), x_3(0), x_4(0), x_5(0), x_6(0)] = [1, 0, 1, 0, 1, 0]$,<br>cancer incidence := having $M_t$ or more malignant cells,<br>$\alpha_3$ is known and $\beta_3$ is not required. |

\* Survival: The probability of having less than $M_t$ malignant cells. For the MSCE model, $M_t = 1$ .
\*\* Hazard rate: the instantaneous rate at which a specific event occurs.
<sup>†</sup> FSMC: First-Stage Mutated Cell, <sup>‡</sup> SSMC: Second-Stage Mutated Cell,
<sup>◊</sup> Aggressiveness: Likelihood of newly proliferated cells to mutate to the next stage.

Suppose *T* is a random variable that denotes the time when a tumor reaches the threshold number of malignant cells for diagnosis (this threshold is assumed to be 1 in the classic MSCE models, and computed based on reported tumor-size-at-diagnosis in the MSCE-T models). We define survival at time *t, Pr*[*t < T*], as the probability of not having a diagnosed tumor by time *t*. If *M*_*t*_ is the threshold malignant cell number at time *t* corresponding to the tumor-size-at-diagnosis and *M* (*t*) is the number of malignant cells at time *t*, then *Pr*[*t < T*] = *Pr*[*M* (*t*) *< M*_*t*_]. If *I*_0_(0), *I*_1_(0), *I*_2_(0) and *M* (0) denote the initial number of normal stem cells, FSMCs, SSMCs, and malignant cells, respectively, then the survival probabilities given different initial conditions are:

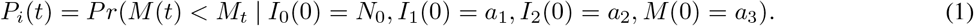

Here, *N*_0_ represents the number of normal stem cells (see Table 1) and (*a*_1_, *a*_2_, *a*_3_) is equal to (0, 0, 0), (1, 0, 0), (0, 1, 0) or (0, 0, 1) when *i* = 0, 1, 2, 3, respectively. These initial conditions are chosen so that the transition probabilities associated with birth, death, and mutation can be written as a product of two or more functions in (1). The age-specific function of cancer incidence or hazard [11] is then:

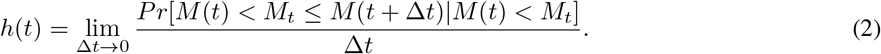

Using the definition of conditional probability and given that *Pr*[*M* (*t*) *< M*_*t*_] = 1 − *Pr*[*M* (*t*) ≥ *M*_*t*_], we can express the hazard function for the survival probability *P*_0_(*t*) as:

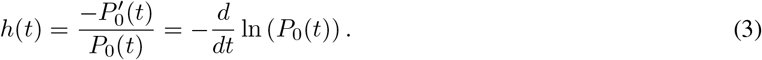

Further, we adopt the following naming conventions:

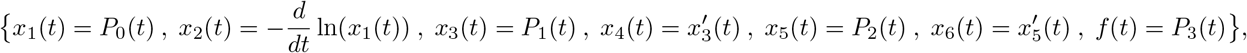

and derive a system of Ordinary Differential Equations (ODEs) to describe the changes in survivals (i.e., the probabilities given in (1)) and the hazard defined by (3):

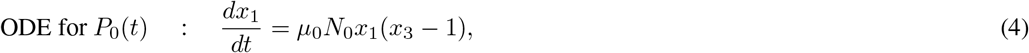

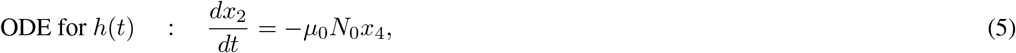

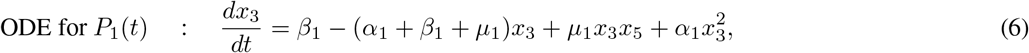

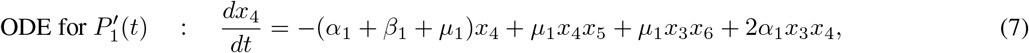

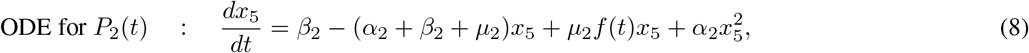

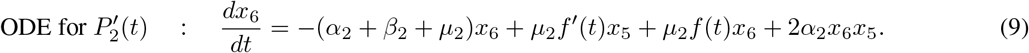

As detailed in the Supplementary Material, the derivation process entails tracking how stem cells transition from normal to malignant as they accumulate genetic mutations. The model considers a birth-death process for each stage, where cells can divide, die, or mutate at specific rates. By incorporating the probabilities of these transitional events, we estimate the probability that a certain number of malignant cells will develop and form a detectable tumor by a given age. Table 1 gives all the variable and parameter descriptions.

In the classic MSCE model, *f* (*t*) = 0, since *M*_*t*_ = 1 and *α*_3_ = *β*_3_ = 0. Here, to incorporate the tumor size, we derive the probability of having less than *M*_*t*_ malignant cells given one malignant cell at *t* = 0 through a linear birth-death process (see Bailey chapter 8 [31]):

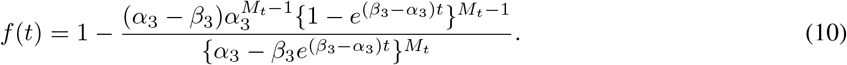

Briefly, to derive equation (10), we first solve the moment-generating partial differential equation of the linear homogeneous birth-death process. From the solution, we then obtain the probability generating function and use that function to generate a general formula for the probability of having exactly *M*_*t*_ malignant cells. Using a finite sum over *M*_*t*_ and the geometric series formula, we arrive at equation (10); see the details in the supplementary material. Combining this function with (4)-(9) gives a general model accounting for tumor-size-at-diagnosis (referred to as the General MSCE-T model in this paper).

A well-accepted simple model of cancer growth is the logistic growth with the rate of 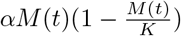 [32], where parameters *α* and *K* are net proliferation rate and carrying capacity, respectively, and *M* (*t*) is the number of cancer cells at a given time *t*. Most diagnosed cancers are still actively growing, indicating the carrying capacity is much larger than *M* (*t*) such that tumor growth prior to detection can be approximated by a simple birth process *αM* (*t*). Hence, by setting *β*_3_ = 0 in equation (10), we obtain the probability of having less than *M*_*t*_ malignant cells assuming a simple birth process:

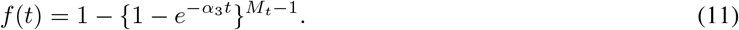

Combining function (11) with equations (4)-(9) leads to a simplified model to account for tumor-size-at-diagnosis and our main MSCE-T model (referred to as the MSCE-T model hereafter). Note that this formulation directly links the time from the occurrence of the first malignant cell to tumor diagnosis via the input *M*_*t*_, as opposed to assuming a constant lag-time in previous MSCE modeling studies. The MSCE-T model can be further simplified using the reported doubling times for different cancer types in the literature, by setting 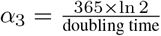 in (11). Here, we use the reported average doubling time of 193 days for BrC [33], 211 days for CRC [34, 35], and 967 days for ThC [36]. This simplification is not feasible for the General MSCE-T model, as *α*_3_ − *β*_3_ (the net proliferation rate) is not uniquely determined, and the term *β*_3_ appears with an exponential factor in the denominator. Thus, we instead use the reported doubling times to establish bounds for estimating *α*_3_ and *β*_3_ in the General MSCE-T model.

Finally, to solve (4)-(9) along with (10) or (11) we consider the initial conditions [*x*_1_(0), *x*_2_(0), *x*_3_(0), *x*_4_(0), *x*_5_(0), *x*_6_(0)] = [1, 0, 1, 0, 1, 0]. However, solving the classic MSCE model (i.e., *f* (*t*) = 0) will entail a different set of initial conditions namely [*x*_1_(0), *x*_2_(0), *x*_3_(0), *x*_4_(0), *x*_5_(0), *x*_6_(0)] = [1, 0, 1, 0, 1, − *µ*_2_]. The initial values for *x*_1_,*x*_3_, and *x*_5_ are one because they represent survival at time zero. The initial values for *x*_2_, *x*_4_ and *x*_6_ are a consequence of their definition (see Table 1) which can be directly determined from equations (4), (6) and (8).

### Model identifiability

A model is structurally identifiable if model parameters can be determined uniquely given perfect (noise- and error-free) data, indicating that there is a unique mechanism as represented by the estimated parameters that can explain the observation. Conversely, a model is termed structurally unidentifiable if several parameter sets yield the same data, indicating unreliable parameter estimation. As such, structural identifiability is important for parameter estimation.

The classic MSCE model, despite its simplicity, is not structurally identifiable. Using a differential algebraic approach, Brouwer et al. prove that for the MSCE model, the system is unidentifiable and the parameters need to be combined into groups to achieve full structural identifiability [39].

The General MSCE-T model does not resolve the identifiability issue. Moreover, due to highly nonlinear terms, including two extra unknown parameters *α*_3_ and *β*_3_, recovering identifiable groups like that of the classic MSCE model is difficult. Please refer to the supplementary material for a justification.

For the MSCE-T model, when *α*_3_ in equation (11) is known (based on values reported in the literature), and *M*_*t*_ is obtained from the data, equation (11) can be treated as a known input to the model. Introducing additional inputs is a recognized approach to mitigate structural unidentifiability, and incorporating (11) instead of (10) makes the model fully structurally identifiable. A detailed proof is provided in the supplementary material.

### Model validation using synthetic data

We validate the model and parameter estimation approach using synthetic data. The synthetic dataset is generated through Poisson sampling, using the model-derived incidence as the mean, to simulate the imperfect observations in real-world settings (see the “ Materials and methods” section for more details). As the parameters are prescribed, we can compare the model estimates with the true parameter values (typically unknown for real-world data) in addition to model fit. Here, we assess i) the model fit to the data, ii) the accuracy of parameter estimation from the best-fit model run, and iii) the consistency based on the ensemble from 100 runs. As shown in Figure 2, the MSCE-T model performs best in all aspects – it affords the best model fit [lowest negative log-likelihood and Akaike Information Criterion (AIC) values], yields the lowest parameter estimation error relative to the true parameter values, and demonstrates high consistency, as evidenced by the tighter distributions of estimated parameters. These findings are consistent with the structural identifiability of the MSCE-T model.

**Figure 2.**
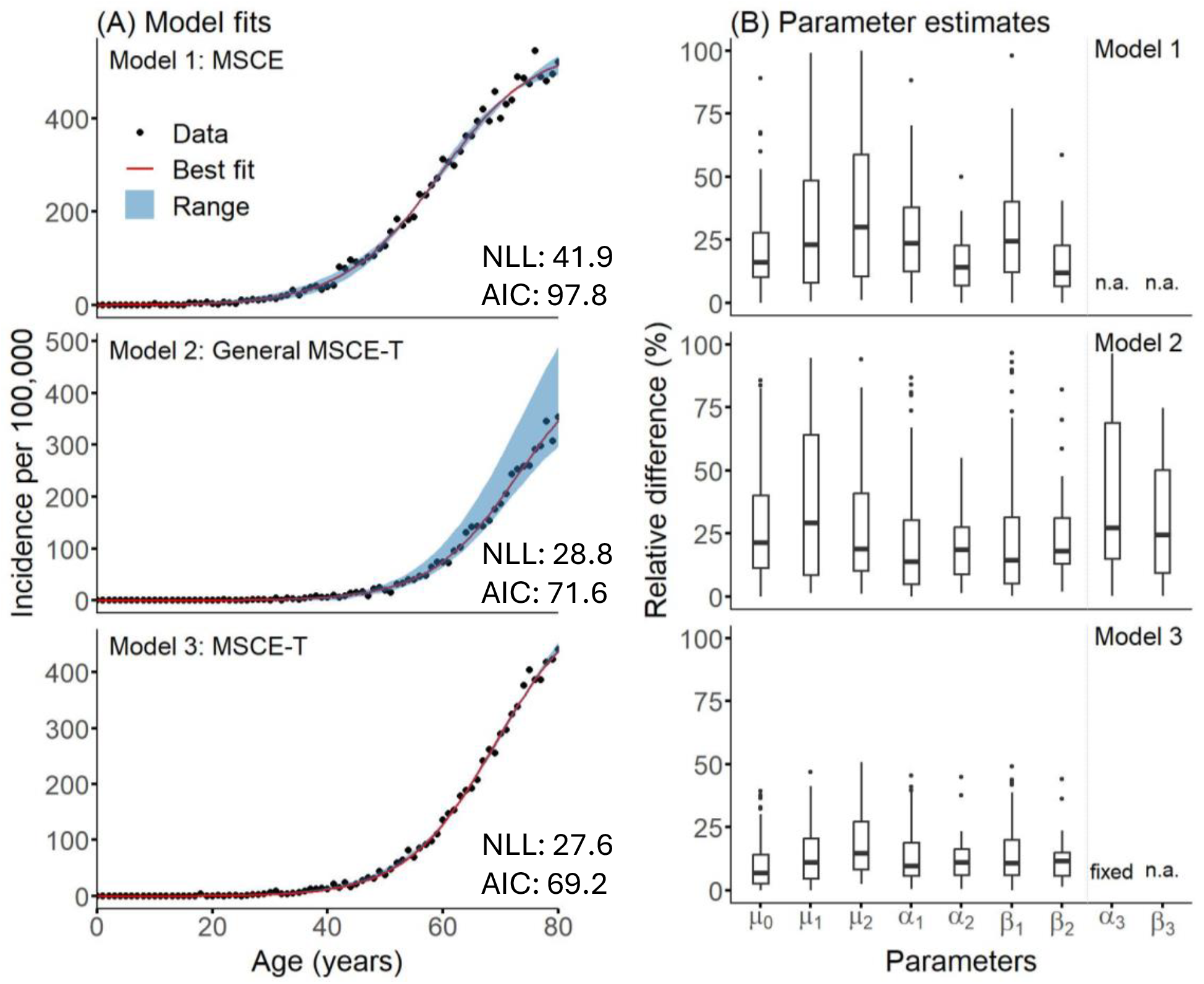
Model validation using synthetic data. (A) Shows the synthetic incidence data (dots) and best model fits (red lines and ranges). (B) Shows the percentage of the relative difference between the estimated parameters and the true values used to generate the synthetic data. The chosen true values for all the models are (*µ*_0_, *µ*_1_, *µ*_2_, *α*_1_, *α*_2_, *β*_1_, *β*_2_) = (5.20 × 10^*−*5^, 3.62 × 10^*−*5^, 1.09 × 10^*−*3^, 2.28, 4.87, 2.16, 4.73). For the General MSCE-T model, we consider *α*_3_ = 9 and *β*_3_ = 6.43. The value of *α*_3_ in the MSCE-T model is fixed and is equal to *α*_3_ − *β*_3_ from the General MSCE-T model. The Negative Log Likelihood (NLL) and Akaike Information Criterion (AIC) are given for the best fits.

## Data analysis results

### Parameter estimates accounting for tumor size

We next apply the MSCE-T model to examine the tumor growth kinetics of three early-onset cancers – BrC (under age 40), CRC (under age 50), and ThC (all ages included), based on cancer incidence across three representative cohorts (1950-1954, 1965-1969, and 1980-1984) in the US. All three cancer types saw increases in incidence among young adults in more recent cohorts (Figure 3; darker colors for more recent cohorts). The MSCE-T model is able to capture the observed incidence trends for all three cohorts (Figure 3). For ease of interpretation, we combine the parameters into three groups representing three key stages of carcinogenesis considered in the model: the mutation rate of normal stem cells (i.e., initial mutation), the FSMC aggressiveness (combining the stage-specific mutation-, birth-, and death rate; Table 1), and the SSMC aggressiveness. Here, aggressiveness can be interpreted as the net rate at which proliferating cells contribute to new mutations at each stage. Importantly, here we computed these parameter combinations directly using individual parameter estimates from the MSCE-T model, given it is fully identifiable, rather than through reparametrization. Figure 4 shows that across the three cancer types, CRC has the highest estimated mutation rate of normal stem cells and the lowest estimated aggressiveness of SSMCs, while estimated aggressiveness for the FSMCs is the highest in ThC.

**Figure 3.**
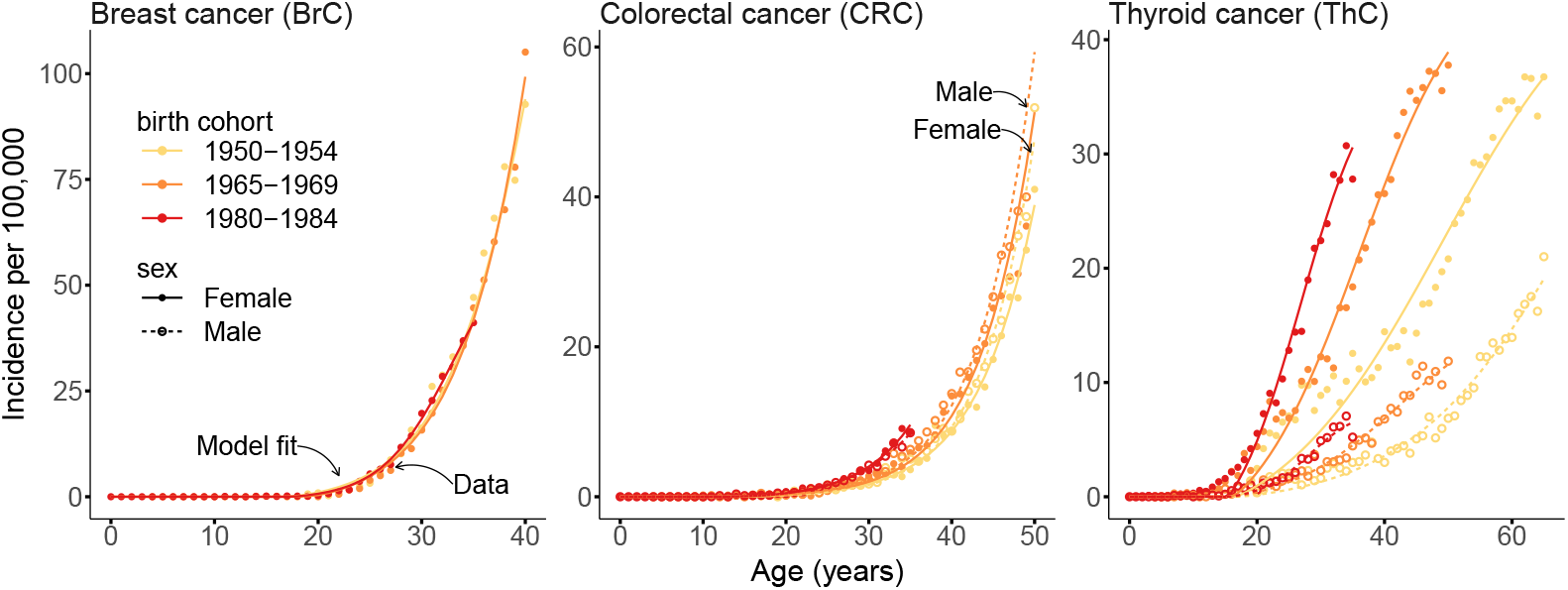
Cancer incidence curves. The curves are produced by fitting the MSCE-T model to incidence data for three different cancer types and three cohorts born in 1950-1954, 1965-1969, and 1980-1984.

**Figure 4.**
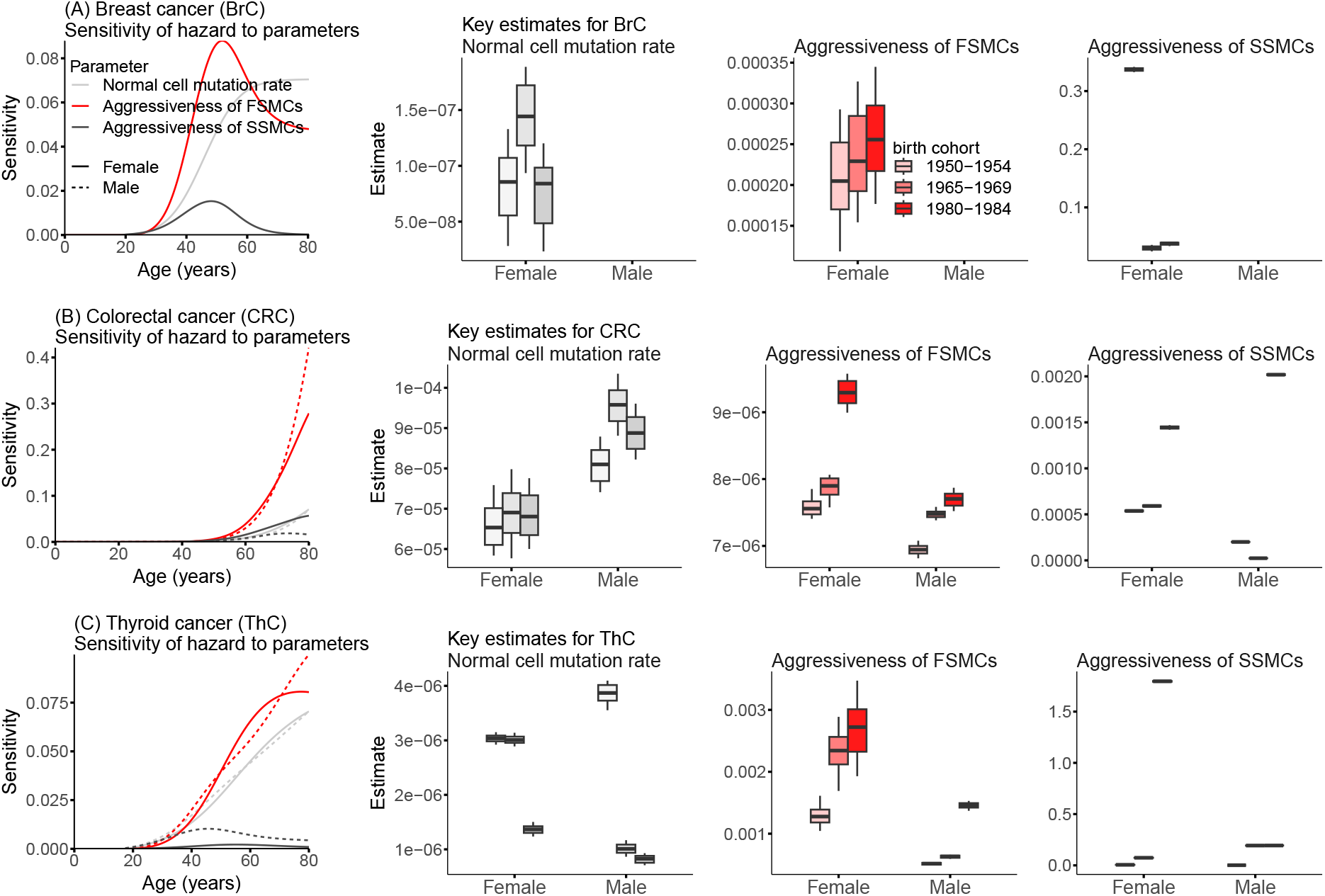
Sensitivity analysis and estimated parameters distribution, for (A) Breast cancer, (B) Colorectal cancer, and (C) Thyroid cancer. The curve plots show the sensitivity of the MSCE-T model hazard to parameters: i) Normal cell mutation rate *µ*_0_; ii) Aggressiveness of the first-stage mutated cells (FSMCs) *µ*_1_ × (*α*_1_ − *β*_1_); and iii) Aggressiveness of the second-stage mutated cells (SSMCs) *µ*_2_ × (*α*_2_ − *β*_2_). The box plots show the distribution of these parameters for three cohorts. Parameters *µ*_*i*_, *α*_*i*_ and *β*_*i*_ all have the unit 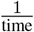. The sensitivity here refers to the unit change in the incidence per unit change in the corresponding parameter value.

For all three cancer types, sensitivity analysis indicates the FSMC aggressiveness [i.e., *µ*_1_ × (*α*_1_ − *β*_1_)] incurs the highest sensitivity such that per-unit change in this parameter combination leads to the largest change in cancer incidence (Figure 4, sensitivity plots, red curves). The MSCE-T model estimates an increase in the FSMC aggressiveness in more recent birth cohorts (Figure 4, second column of box plots, higher red bars in later cohorts) for all three early-onset cancers. Particularly, for female BrC, compared to women born in 1950-1954, the estimated FSMC aggressiveness shows increases of 13.6% (95% CI: 6-21; 1965-1969 cohort) and 23.2% (95% CI: 16-31; 1980-1984 cohort) in a span of 30 years. For CRC, compared to the 1950-1954 birth cohort, estimated FSMC aggressiveness increased by 3.9% (95% CI: 0.1-9; female) and 7.8% (95% CI: 5-10; male) in the 1965-1969 birth cohort and by 22.2% (95% CI: 16-29; female) and 10.4% (95% CI: 7-14; male) in the 1980-1984 birth cohort. For ThC, the estimates increased by 80.7% (95% CI: 20-147; female) and 22.2% (95% CI: 12-32; male) in the 1965-1969 birth cohort and by 111.9% (95% CI: 32-191; female) and 180.9% (95% CI: 158-206; male) in the 1980-1984 birth cohort. In addition, the model estimates higher values for FSMC aggressiveness in females than males for both CRC and ThC (Figure 4, see the second column of box plots, higher darker red bars for females). Estimates for the other two parameter combinations are less consistent across cohorts and sexes, and as noted, the model is less sensitive to these parameter combinations.

### Sensitivity and supplemental analyses

To assess the robustness of our model estimates, we conducted two sensitivity analyses: 1) we varied the values of *α*_3_ for all three cancer types, given that this parameter is derived from the literature; and 2) we set the tumor-size distribution for years before 1988 (no available data) to the reported distribution in 1988 (i.e., the earliest year with data), as opposed to extrapolating the tumor-size-at-diagnosis data in the main analysis (see Materials and Methods). Figure S2 compares parameter estimation results for different settings of *α*_3_, and Figure S3 compares results for different handling of missing tumor size data. Despite the variation in parameter values, for both sensitivity analyses, the qualitative trends remain consistent with the results reported above (and shown in Figure 4) across all parameter combinations and cancer types. Particularly, the most sensitive parameter (i.e., the aggressiveness of FSMC) consistently increases by birth cohort in all analyses.

As noted in the Introduction, improvement in cancer detection and incidental detection due to increased use of medical imaging could contribute to the apparent increases in cancer incidence. Such biases in the data are more profound for ThC (see the large increases in Figure 3) 7]. To examine the ability of the MSCE-T model to account for such detection-related data biases, we conducted two additional analyses. First, we compared parameter estimates using the MSCE model, which does not account for tumor size, with estimates from the MSCE-T model. The MSCE model yields inferior model fit (see Table S4) and its parameter estimates do not show clear changes by birth cohort for the three cancer types examined (Figure S4). In the second analysis, we performed parameter estimation for ThC by subtype. About 90% of ThC cases are papillary carcinomas, 1% are anaplastic carcinomas, and the remaining cases are follicular, Hürthle, and medullary carcinomas [40]. Despite its scarcity, anaplastic ThC accounts for over 30% of all ThC-related deaths [40]. Thus, we conducted the analysis for papillary (most prevalent) and anaplastic (most lethal) ThC separately. Given the low incidence of anaplastic ThC, for this analysis, we aggregated the data for 15-year cohorts (i.e.,1940-1954, 1955-1969, and 1970-1984) to reduce observational noise. However, we note that the mean incidence of anaplastic ThC is still low (Figure S5) and caution the greater uncertainty in model estimates for this subtype. As shown in Figure S5 (A), the MSCE-T model estimates increases in the FSMC aggressiveness for papillary ThC over the three birth cohorts, similar to the main analysis combining all ThC subtypes. In contrast, for anaplastic ThC, there are no clear changes in incidence over the three cohorts for both sexes, and the model does not estimate an increase in FSMC aggressiveness (Figure S5 (B)). Together, these results demonstrate the ability of the MSCE-T model to account for changing detection and identify changes in tumor growth kinetics and suggest there are increases in the risk of the three early-onset cancers (for ThC, such increases are mostly related to papillary ThC) in more recent cohorts, independent of changing detection.

### Sojourn time

We use the MSCE-T model to calculate the mean sojourn time (i.e., the average time from the emergence of first malignant cell to cancer detection) of BrC, CRC, and ThC for the three cohorts (1950-1954, 1965-1969, and 1980-1984). An illustration of this calculation is provided in Figure S6. The mean sojourn times (Table 2) estimated here are comparable to results from the Cancer Intervention and Surveillance Modeling Network (CISNET), for example, 2-4 years for breast cancer [41] and 10.6 years (with an interquartile range of 5-14 years) from adenoma incidence to cancer diagnosis for CRC [42]. Here, we provide more detailed estimates by sex and birth cohort. The estimates are similar for females and males of the same cohort. However, we notice a decreasing trend for more recent cohorts for all cancer types. To assess the influence of the likely improved detection over time on the estimates, we fixed *M*_*t*_ at the mean across all cohorts to compute the sojourn times controlling for changes in tumor-size-at-diagnosis. The declining trend in sojourn time persisted but with a slight reduction (though more noticeable for ThC) in each estimate. Together, these results suggest increases in tumor aggressiveness (hence the shorter sojourn times), consistent with the model parameter estimates reported above.

**Table 2.** Mean sojourn time for different cancer types and cohorts.

| Cancer Type | Sex | Cohort (Years) | Mean sojourn time with the actual $M_t$ (Years) | Mean sojourn time with fixed $M_t^*$ (Years) |
| --- | --- | --- | --- | --- |
| BrC | Female | 1950-1954 | 4.2 (CI:3.9-4.4) | 4.1 (CI:3.8-4.4) |
|  |  | 1965-1969 | 4.1 (CI:3.8-4.4) | 3.9 (CI:3.8-4.4) |
|  |  | 1980-1984 | 4.0 (CI:3.7-4.3) | 3.8 (CI:3.5-4.0) |
| CRC | Female | 1950-1954 | 10.7 (CI:9.4-12.0) | 10.5 (CI:9.1-11.8) |
|  |  | 1965-1969 | 10.1 (CI:9.2-11.1) | 10.0 (CI:9.1-10.9) |
|  |  | 1980-1984 | 8.8 (CI:7.6-9.8) | 8.7 (CI:7.6-9.8) |
| CRC | Male | 1950-1954 | 10.7 (CI:9.3-12.0) | 10.5 (CI:9.2-11.9) |
|  |  | 1965-1969 | 10.1 (CI:9.2-11.0) | 10.0 (CI:9.1-11.0) |
|  |  | 1980-1984 | 8.8 (CI:7.6-9.8) | 8.8 (CI:7.7-9.9) |
| ThC | Female | 1950-1954 | 17.1 (CI:16.8-17.3) | 16.3 (CI:15.4-17.4) |
|  |  | 1965-1969 | 14.7 (CI:14.2-15.3) | 14.4 (CI:13.9-15.0) |
|  |  | 1980-1984 | 12.3 (CI:11.9-12.7) | 12.2 (CI:11.6-12.8) |
| ThC | Male | 1950-1954 | 17.0 (CI:16.7-17.2) | 16.4 (CI:15.5-17.2) |
|  |  | 1965-1969 | 14.7 (CI:14.2-15.2) | 14.5 (CI:14.1-14.8) |
|  |  | 1980-1984 | 12.2 (CI:12.0-12.5) | 12 (CI:11.3-12.8) |

\* $M_t$ is fixed as the average tumor cell counts at diagnosis across all cohorts.

### Impact of population screening on cancer incidence

Given the mechanistic design, the MSCE-T model also affords prediction to help assess the impact of population screening on cancer incidence. Particularly, for BrC and CRC, the parameters are estimated using incidence data before the recommended screening ages. These estimates thus represent tumor growth kinetics without screening, and the model projections would represent incidence under a counterfactual scenario where there is no population screening. To test model accuracy, we first use these parameter estimates to project the incidence of BrC for ages above 40 and CRC for ages above 50 in an early birth cohort with limited screening (1945-1949; i.e., “ control” cohort). Indeed, as shown in Figure 5 first panel, for both BrC and CRC, the model-projected incidence rates roughly match the observed data for this “ control” cohort (<30% above the age of 40 screened for BrC and <34% above the age of 50 screened for CRC [44]). These results indicate the model is able to sensibly capture BrC and CRC risk in the absence of screening.

**Figure 5.**
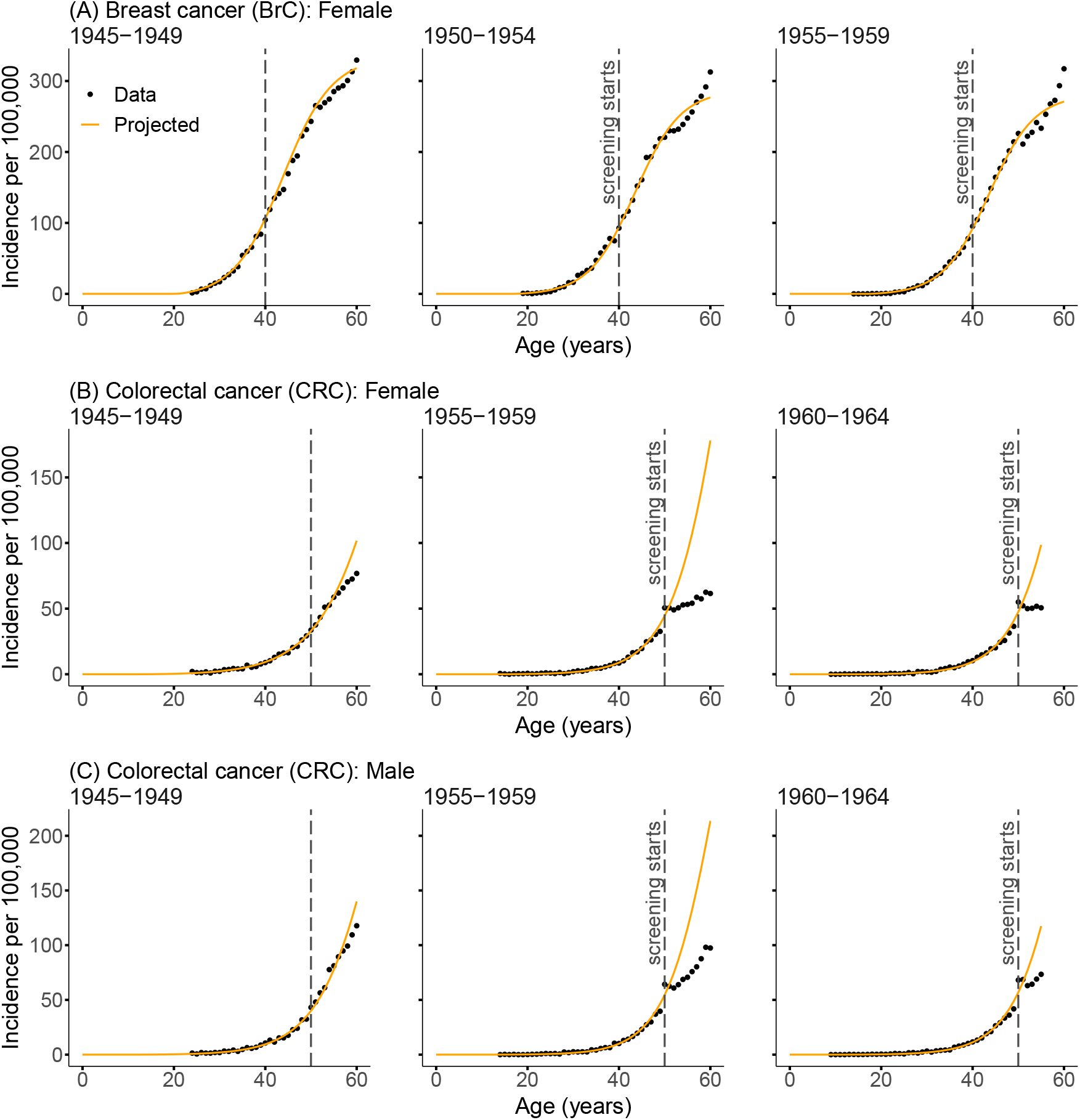
Counterfactual modeling to examine the impact of population-level screening for BrC and CRC. The MSCE-T model is fitted to female BrC patients born in 1945-1949, 1950-1954, and 1955-1959 (sub-figure A). For CRC, the model is fitted to patients born in 1945-1949, 1955-1959, and 1960-1964 (sub-figures B for female and C for male patients). The dashed lines mark the screening initiation. The model is first fitted to incidence before the recommended screening age (dots to the left of the dashed lines) and then used to generate projections (orange curves; i.e., counterfactuals with no screening) for older ages, compared to the actual incidence (dots to the right of the dashed lines). The dashed lines for the cohort of 1945-1949 are not annotated since screening was not yet as ubiquitous for this cohort.

We then assess the impact of screening for these two cancer types in two cohorts with high rates of screening (i.e., the 1950-1954 and 1955-1959 cohorts for BrC and the 1955-1959 and 1960-1964 cohorts for CRC; >~ 60% of eligible population screened). For BrC, model projections for the two screening-affected cohorts (1950-1954 and 1955-1959) align closely with the observed incidence until approximately ages 50-55 (Figures 5 (A) middle and right) – note around this age range, there was a brief transition phase (i.e., slow-down) in incidence rates, a phenomenon termed Clemmensen’s hook [45], which was not accounted for in our model projections. Around ages 56-60, the observed incidence rates start to more substantially surpass the model projections (see, e.g., Figure 5 (A) middle vs left; dots above the lines). This larger discrepancy between the model projection and the data for these cohorts likely reflects the increased detection of BrC through screening, consistent with the higher percentage of screen-detected than interval-detected cases reported for ages ≥ 60 [46]. Nonetheless, as a slight discrepancy is also seen in the “ control” cohort with limited screening, we suspect the higher-than-projected incidence is in part due to a true risk increase in those ages as noted in [45].

For CRC, among those aged 50 and above, projected incidence (without screening) is much higher than the observed incidence for the two screening-affected cohorts (1955-1959 and 1960-1964) for both females and males (Figures 5 (B) & (C) middle and right). This discrepancy likely reflects the preventive impact of CRC screening using colonoscopy, which not only detects but also facilitates the removal of precancerous polyps, thereby preventing malignancies from progressing to clinically observable cases [47].

## Discussion

This study introduces an extension to the classic multistage carcinogenesis model by including tumor-size-at-diagnosis data. Our findings suggest that adding this input significantly improves the model’s sensitivity and ability to capture important trends in cancer progression. The MSCE-T model outperformed the classic MSCE model in terms of structural identifiability and parameter estimation.

To gauge model accuracy, we compare the mean sojourn times estimated using our model with independent estimates from the literature. Luebeck et al. (2013) used a similar routine and definition for sojourn time as in this study (i.e., from the occurrence of the first malignant cell to cancer diagnosis). However, their estimates – 5.2 years (CI: 3.6–6.2) for male CRC and 6.5 years (CI: 5.2–7.6) for female CRC [20] – are much shorter than our estimates (8-10 years in Table 2). Two main methodological differences may have contributed to this discrepancy. First, Luebeck et al. (2013) treated detection as a stochastic event, whereas our model explicitly tracks tumor growth from the first malignant cell to diagnosis based on reported tumor size data. Second, Luebeck et al. (2013) adjusted their incidence data for cohort and period effects, while we used the cohort-specific data directly. The CISNET MISCAN-Colon output a mean sojourn time of 10.6 years (CI: 5-14) from adenoma incidence to CRC diagnosis [42], although there is large uncertainty in the time spent in the adenoma stage and the probability of progressing to CRC [42]. The CISNET-DFCI model estimated a sojourn time of 2–4 years for BrC in the pre-screening era [41]). These model estimates are close to ours (Table 2). However, we caution the differences in methodology and assumptions - CISNET models rely on simulated populations with assumptions about transition dynamics or treat sojourn time as a calibrated input [42, 41], whereas we estimate these durations using the MSCE-T model, incorporating cancer incidence and tumor-size-at-diagnosis data.

For ThC, we estimated much longer sojourn times (~ 15 years; Table 2), despite the earlier lifetime occurrence of ThC than the other two cancer types (i.e., BrC and CRC; see age-specific incidence in Figure 3). Given the elevated incidence rates starting around ages 15-20 (Figure 3), these estimates suggest that the initiation of ThC might have started at a very early stage of life. We are unable to locate a similar model estimation of sojourn time for ThC for comparison (it is less modeled). However, extensive clinical studies have also pointed to the likely initiation of ThC during the infantile period [48]. Particularly, studies of children and adolescents exposed to the Chernobyl nuclear accident reported the highest incidence rate of ThC among those who were under 1 year of age at the time of Chernobyl, and the rates decreased progressively through age 12 [49]. Large population-scale health surveys of Fukushima residents, conducted to monitor the impacts of the TEPCO-Fukushima Daiichi Nuclear Power Plant accident, also revealed high baseline (i.e., not associated with the accident) incidence rates of ThC among children, particularly those aged 15 and older [50, 51]. Further, detailed autopsy data showed the prevalence of ThC (i.e., identified through autopsies of people who died from mostly non-ThC-related causes) increases steeply from age 15 to 34 and then stays roughly constant for the remaining lifetime (see Figure 2 of Takano 2017 [48]). This is possible as ThC is self-limiting (i.e., malignant yet causing very low mortality; [48]). The consistency of our model estimates with these independent clinical observations indicates that our model is able to accurately identify the underlying cancer kinetics.

The changes in the kinetics of early-onset cancers are particularly of interest, given the dramatic increases in incidence during recent years in the US and globally [2]. After controlling for potential data biases due to changing detection, estimated carcinogenic aggressiveness still increased substantially for the three studied cancers, i.e., BrC, CRC, and ThC, which indicates a genuine rise in the underlying cancer risk. We identified the aggressiveness of the first-stage mutated cells (i.e., FSMC aggressiveness) to be the most impactful intermediate step affecting cancer risk. Here, aggressiveness reflects both a cell population’s mutagenic potential and its capacity for clonal expansion, which could contribute to the formation of a cancerized field - namely, a population of cells that, while not fully malignant, have acquired early genetic alterations that predispose them to neoplastic progression [52]. Such a cancerized field can serve as a substrate with enhanced expansion capacity, effectively increasing the likelihood of tumor development. Across the 30-year study period, we estimated FSMC aggressiveness rose significantly for both sexes (e.g., ~ 23% increase in female BrC; ~ 10% in male CRC; see Figure 4), with consistently higher estimates in females. Furthermore, based on the recent study by Gerstung et al. [53], we hypothesize that the estimated increase in carcinogenic events — reflected in the higher aggressiveness of FSMC — likely arises during early to mid-adulthood for the more recent cohorts (e.g., those born during 1980-1984). A more detailed characterization of the timing of gene-specific mutations, building on the model framework developed here, represents an opportunity for future research.

Early detection and treatment through screening have been a key cancer intervention strategy. In the US, population screening is recommended for both BrC (age ≥ 40) and CRC (age ≥ 50, or ≥ 45 from 2021 onward [5]). As clinical trials are difficult to conduct in younger ages when cancer is rare, modeling often serves as a means to evaluate the effectiveness of screening as well as etiology [54]. In particular, several large-scale CISNET models have been developed and applied to test different screening policies and their effectiveness for both cancer types [55, 56, 57, 58]. Here, our model provides an alternative approach to assess the effectiveness. Using counterfactual modeling, we show striking reductions in CRC incidence among those aged ≥ 50 (Figure 5 (B) & (C) middle and right), likely thanks to the implemented screening programs and the feasibility of removal of precancerous polyps detected during screening [47]. For BrC, our modeling analysis shows no decline in BrC incidence following screening and, instead, higher BrC incidence for those aged ≥ 56 − 60, likely detected through screening. Such differences compared to CRC likely arise from a combination of factors. For one, breast lesion removal is much more complex [35]. For another, BrC risk varies nonlinearly around ages 45-55 (Clemmensen’s hook [45]); note again our model did not account for such changes. Nonetheless, our model projections for BrC are consistent with observations [46] - that more BrC cases in women under age 50 were interval-detected than screen-detected (hence the projected incidence without screening and the observed incidence with screening are similar for those under 50) and that more cases in women aged ≥ 56 − 60 were screen-detected (hence higher observed BrC incidence than the projections). Further, unlike the CISNET models assessing screening impact primarily at the population level without explicitly accounting for cohort-specific trends [59, 60, 42, 61], our model accounts for the apparent incidence changes due to changing detection and constructs cohort-specific counterfactuals, based on cohort- and age-specific incidence and tumor-size-at-diagnosis data that are readily available from the SEER program. This simplicity and specificity of our model thus affords a powerful alternative to more explicitly assess the screening impacts, particularly in the context of evolving early-onset cancer risk.

We recognize several study limitations. First, based on findings from Tomasetti et al. [25] and for generality, we adopt a three-stage model for all three cancers studied here. While the model estimates are consistent with the literature, as noted in the Results, we cannot determine whether three rate-limiting mutations indeed apply to the three cancers (i.e., BrC, CRC, and ThC). Second, while we are able to estimate the changes in cancer kinetics, the current model does not consider the underlying causes for the estimated changes. Future work can incorporate risk factor data (e.g., as done in [21]) to help identify the main underlying causes. Third, for simplicity, we assume constant values for the parameters in this study. Time-dependent parameters (e.g., based on risk factor data) would likely further improve model performance and provide more detailed estimates, particularly the likely causes of the increased cancer risks during different intermediate stages of tumor growth. Fourth, the tumor mass is likely highly complex, encompassing various cell types. Here, for simplicity, we did not consider such heterogeneity when converting the tumor size to the number of malignant cells (i.e., *M*_*t*_ in the model). Nonetheless, we note this simplification would not affect the estimated changes by birth cohort, as shown in our analysis of mean sojourn time setting *M*_*t*_ to a fixed number for all cohorts; Table 2. Fifth, the current model captures period effects related to diagnostic practices through tumor-size-at-diagnosis data, but sidesteps other non-detection-related period effects which could potentially influence cancer incidence. Finally, for simplicity, we did not include stochasticity reflecting the individual-level heterogeneity in data, which may in turn underestimate the parameter uncertainties. We intend to improve this model and its applications by addressing these limitations in our future work.

In conclusion, this study introduces an improvement in cancer modeling by integrating tumor-size-at-diagnosis data into the multistage carcinogenesis framework. This approach improves model accuracy and ability to detect subtle trends in cancer risk by controlling for observational biases due to changing detections. Despite the noted limitations, the proposed MSCE-T model offers valuable insights into tumor growth kinetics, particularly for early-onset cancers that have increased in recent years. In addition, the model provides an alternative approach to evaluating the impact of population screening programs, as illustrated here for breast and colorectal cancers. These findings could inform early cancer prevention programs.

## Materials and methods

### Data sources and processing

We use the SEER cancer incidence registries for patients diagnosed during 1973-2015 with BrC, CRC, and ThC. We use the International Classification of Diseases for Oncology (ICD-O) codes to filter out the three cancer types: 1) C50.0-C50.9 for BrC; 2) C180.0-C180.9, C19.9, and C20.9 for CRC to include the colon, rectosigmoid junction, and rectum; and 3) C73.9 for ThC. We divide the data into 5-year birth cohorts and stratify them by sex.

The SEER dataset reports the tumor-size-at-diagnosis for 1988-2015, while the year of diagnosis goes as far back as 1973. The size data are reported under 10-digit EOD (1988-2003) or CS tumor size (2004-2015) and describe the largest dimension, or the diameter of the primary tumor, at the time of diagnosis. Using linear extrapolation, we estimate tumor size at detection for years without SEER data (1973-1987); see Figure S1 in the supplementary material. Figure 1 (A) shows the average size data extracted from the SEER registry. A decreasing trend is evident for all three cancer types. We use a formula based on Kepler’s conjecture to compute the number of cells from the tumor size (see details in the supplementary material).

Normal mammary stem cells are in a quiescent state (i.e., inactive) until puberty [62]. Hence, for all analyses related to BrC, we solve the ODEs with a delay corresponding to age at menarche for each cohort reported in Table S3. Data on age at menarche were collected by the National Health and Nutrition Examination Survey (NHANES) using reproductive health questionnaires [63]. We pooled all data from the 1999-2000 to 2017-2018 cycles and adjusted sample weights across survey cycles following NHANES guidance. After restricting respondents to those born between 1930 and 1994, the response ranged from 8 to 20 years, 21,830 individual responses remained in this analysis. We then computed age at menarche (mean and 95% CI) for each 5-year birth cohort (i.e., those born during 1930-1934, …, 1990-1994), accounting for sample weights using the “ survey” package in R [64].

### Model validation using synthetic data

We compare the parameter estimation for each of the three models. Models MSCE and MSCE-T each have seven parameters, and the General MSCE-T model, with the unknown *α*_3_ and *β*_3_, has nine parameters. To make the comparison fair, we add a constraint for this model’s parameter estimation, forcing the algorithm to find *α*_3_ and *β*_3_ values such that *α*_3_ − *β*_3_ is equal to the pure birth rate of the MSCE-T model. We generate synthetic data for the incidence curves using the models and an arbitrary parameter set - as the true parameter values are known here, we can assess the accuracy of model estimates directly. To mimic noise in observations, we use Poisson random sampling with the mean set to the model-simulated incidence. We use Hybrid Genetic Algorithm (HGA) optimization for parameter estimation [65], employing MATLAB’s HGA from the Global Optimization Toolbox. We run the algorithm 100 times and record the fittest set of parameters and their fitness value in each iteration. The fitness value is the least square distance of the model output from the data (cost). We consider the fittest (i.e., lowest least square distance) of the 100 parameter sets as the best-fit parameter set. For illustration purposes, we calculate and plot the distribution of the relative difference percentage between the best-fit parameter set and the true value, see Figure 2.

### Parameter estimation for BrC, CRC, and ThC incidence data

We estimate parameters to identify trends in cancer progression across different cohorts and cancer types. In this study, we estimated parameters for BrC, CRC, and ThC based on the data for cohorts born in 1950-1954, 1965-1969, and 1980-1984. These cohorts were chosen to represent distinct historical periods, allowing us to capture temporal trends in cancer biology and treatment advancements. Additionally, these cohorts provide a significant number of early-onset incident cases. To minimize bias introduced by cancer screening for BrC and CRC, we restrict our analysis to incident cases diagnosed before age 40 and 50, respectively. For BrC, as mentioned in the “ Data sources and processing” section, we impose a time delay corresponding to age at menarche for each cohort reported in Table S3. For ThC, since there is no screening age policy, we can use all the available data for these cohorts. As in the synthetic testing, parameter estimation for each cohort is carried out 100 times using MATLAB’s HGA toolbox. We acquire a distribution of the parameter values from 100 iterations. We summarize the results via three biologically meaningful combinations containing all the model-estimated parameters. These combinations represent the three key stages of carcinogenesis considered in the model: the initial mutation rate of normal stem cells *µ*_0_ and the aggressiveness of the 1st and 2nd stage mutated cells, *µ*_1_ × (*α*_1_ − *β*_1_) and *µ*_2_ × (*α*_2_ − *β*_2_), respectively (Table 1). We applied the delta method to estimate confidence intervals for the percentage changes in parameter estimates across birth cohorts.

### Sojourn Time

To calculate the mean sojourn time, first, we acquire an estimation for parameter values by fitting the MSCE-T model to the incidence of CRC (under age 50), BrC (under age 40), and ThC (all ages) for cohorts 1950-1954, 1965-1969, and 1980-1984, as explained in the previous section. Note that this fitting is done considering the tumor cell count at diagnosis (*M*_*t*_). Using the same parameters in the model but setting *M*_*t*_ = 1 will result in cancer incidence being recorded at the first malignancy. Hence, to find the sojourn time, we calculate the difference between the time the MSCE-T model with varying *M*_*t*_ *>* 1 produces the same incidence as the MSCE-T model with *M*_*t*_ = 1. We obtain the mean sojourn time by averaging the sojourn times restricted to cases for which relevant clinical data are available. We repeat the procedure by fixing *M*_*t*_ (average over all cohorts) to explore the effect of detection improvement.

### Impact of population screening on BrC and CRC incidence: a counterfactual analysis

As noted above, we estimate model parameters for ages under 40 for BrC and 50 for CRC, who were not subject to population screening for these cancers during the study period. To examine the impact of population BrC and CRC screening, we then use these parameter estimates and the MSCE-T model to project cancer incidence for those older than the recommended screening ages. That is, as the parameters do not include the impact of screening, these projections represent cancer incidence under a counterfactual scenario with no screening. For female BrC, according to the CDC National Center for Health Statistics, mammogram screening for women over the age of 40 reached 59.7% by 1993 and increased to over 70% by 2000 [43]. Therefore, for analysis of BrC screening, we examine cohorts of 1950-1954 (reaching age 40 in 1990-1994) and 1955-1959 (reaching age 40 in 1995-1999). For CRC, about 59-62% of adults over the age of 50 underwent some screening procedures between 2005 and 2010 [44]. Therefore, for analysis of CRC screening, we examine cohorts of 1955-1959 (reaching age 50 in 2005-2009) and 1960-1964 (reaching age 50 in 2010-2015). These cohorts contain a significant number of data points before and after the screening age. Moreover, for model validation, for both cancer types, we include the cohort of 1945-1949 (less than 30% of the population for BrC [43] and 34% for CRC [44] were screened) to examine accuracy of model projections when screening was much more scarce.

## Data Availability

All data produced in the present study are available upon reasonable request to the authors.

## Author Contributions

NM and WY conceived the study, performed the analysis, and wrote the first draft. CH, MBT, and PD contributed to the study design and interpretation of the results. All authors contributed to the final draft.

## Funding information

This work was supported by the National Institutes of Health (R01CA257971).

## Acknowledgement

We would like to thank The Mailman School of Public Health and The Center for Computational Biology and Bioinformatics (C2B2) at Columbia University for access to high-performance computing resources.

## Competing interest statement

Piero Dalerba is listed as a co-inventor on patents owned by the University of Michigan (US-07723112), Stanford University (US-09329170, US-09850483, US-10344094, US-11130813) and Columbia University (US-12115140-B2), and related to: 1) the discovery of surface markers for the differential purification of cancer stem cell populations from human malignancies; 2) the use of single-cell genomics technologies for the identification of pharmacological targets expressed in cancer stem cell populations; 3) the combination of anti-CD47 and anti-EGFR monoclonal antibodies for the treatment of human colon cancer; and 4) the use of inverse agonists of RAR/RXR signaling as anti-tumor agents against adenoid cystic carcinomas. Some of the patents listed above were licensed to pharmaceutical companies, resulting in the award of royalties and/or stock from Oncomed Pharmaceuticals, Quanticel Pharmaceuticals and Forty Seven Inc. (Gilead). Piero Dalerba recently owned stock of Eli Lilly and Company. Piero Dalerba’s spouse is employed by Regeneron Pharmaceuticals Inc., and owns (or recently owned) stock of the following pharmaceutical companies: AbbVie, Amgen, AstraZeneca, Eli Lilly and Company, Gilead Sciences Inc., GlaxoSmithKline (GSK), Johnson & Johnson, Merck & Co., Novartis, Organon & Co., Pfizer, Teva Pharmaceutical Industries Ltd and Viatris. Chin Hur has served as a consultant for Guardant Health. Other authors declare no potential conflicts of interest.

## Notes

### Author Declarations

The study used ONLY openly available cancer incidence data originally located in the SEER registry dataset.

### Summary of Updates

Major changes in breast cancer analyses considering age at menarche. Added sensitivity analysis for tumor-size-at-diagnosis input. Added details for more clarity.

