## Supplementary material for "Modeling early-onset cancer kinetics to study changes in underlying risk, detection, and impact of population screening"

### Estimating the number of tumor cells

We extract the tumor-size-at-diagnosis from the SEER data. Given the wide distribution, we restrict the analysis to sizes falling within the first and third quartiles. After that, we use a backward-in-time linear extrapolation to estimate the size for earlier years. That is, we estimate the earlier tumor sizes using a normal distribution sampling with the regression line (size $\sim$ year) based on years with available data (1988-2015) as the mean and the calculated deviation as the standard deviation. It is worth noting that even within the years for which tumor size data is available (1988–2015), some individual cases lack this information in the SEER registry. However, such instances are in the minority, with the percentage of available size data increasing over time. Figure S1 illustrates the reported average size, our estimates, and data completeness (i.e., the percentage of cases with tumor size data reported from 1988 to 2015).

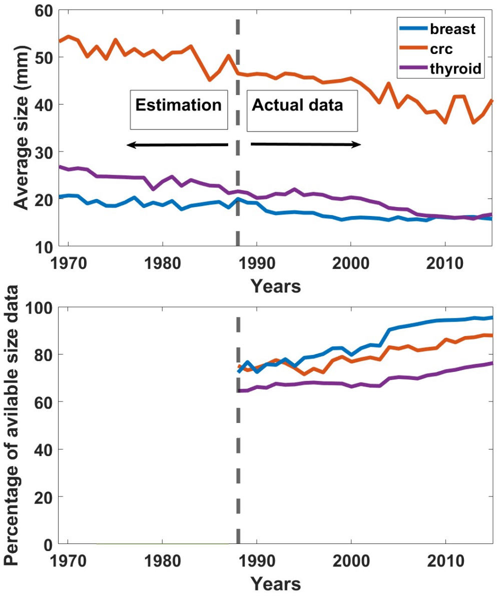


Figure S1: Average tumor size and percentage of cases with reported size data. Top: The mean tumor-size-at-diagnosis for BrC, CRC, and ThC. The data to the right of the dashed line are reported from the SEER data; the data to the left are estimated. Bottom: Percentage of available size data from the SEER registry (1988-2015) for BrC, CRC, and ThC.

As the models estimate the number of malignant cells, we convert the size of the tumor to cell numbers as follows. The reported size describes the longest dimension, also known as the diameter of the tumor. For simplicity, we assume that the tumor's dimension is $R$ in each direction. So, it can be contained in a cube with volume $R^3$. We can also calculate the volume of a single tumor cell, assuming they are nearly spherical. Table S1 shows the reported cell diameters for the tumor cells of interest in this study. Kepler's conjecture claims that when packing $N$ spheres of radius $r$ in the most efficient manner within a face-centered cubic arrangement, the maximum achievable density is approximately 0.74 [1]. The density here is defined as the total volume of spheres divided by the volume of the cube. So, to approximate how many cells of radius $r$ can be packed within a cube of the volume $R^3$, we use the following formula.

$$\text{Cell \#} = 0.74 \times \frac{R^3}{\frac{4}{3}\pi r^3}.$$

Table S1: Tumor cell diameters for three different cancer types.

| Tumor type | Cell diameter (microns) | Reference |
|---|---|---|
| 1. Breast | 31.85 | [2] |
| 2. Colorectal | 7.5 | [3] |
| 3. Thyroid | 10 | [4] |

### Model derivation

The extended model derivation is very similar to the classic model, whose steps have been sketched out by other authors before [5, 6, 7]. Since we want to connect the classic model with the clonal expansion of malignant cells, we reiterate the process from the beginning. To understand the derivation fully, the readers require basic knowledge of stochastic processes, random variables, Markov chains, and Markov properties. In what follows, we summarize the derivation process, assuming that the malignant cells undergo a simple birth-death process. This is the most general form of the model presented in the main text (i.e., the General MSCE-T model). Then, to get the classic model, we set both birth and death rates of the malignant cells equal to zero, and for the malignant cells to undergo a simple birth process, we set their death rate equal to zero.

In this paper, survival is synonymous with the probability of the malignant cells' size being less than the reported size at diagnosis at a given time (age). As mentioned in the main text, we can roughly approximate how much space several malignant cells occupy. So from now on, instead of size, we use the number of malignant cells. Based on a three-stage mutation model, we define the following time-dependent probability functions:

$$P_0(t) = Pr(M(t) < M_t \mid I_0(0) = N_0, I_1(0) = 0, I_2(0) = 0, M(0) = 0), \quad (1)$$
$$P_1(t) = Pr(M(t) < M_t \mid I_0(0) = N_0, I_1(0) = 1, I_2(0) = 0, M(0) = 0), \quad (2)$$
$$P_2(t) = Pr(M(t) < M_t \mid I_0(0) = N_0, I_1(0) = 0, I_2(0) = 1, M(0) = 0), \quad (3)$$
$$P_3(t) = Pr(M(t) < M_t \mid I_0(0) = N_0, I_1(0) = 0, I_2(0) = 0, M(0) = 1). \quad (4)$$

Where $I_0(t), I_1(t), I_2(t)$ and $M(t)$ are random variables denoting the number of normal, first-stage mutated cells (FSMCs), second-stage mutated cells (SSMCs), and malignant cells at time $t$. The value $M_t \neq 0$ is the approximate number of malignant cells occupying a space equivalent to the tumor-size-at-diagnosis reported by SEER data.

According to the multistage clonal expansion biological scheme provided in the main text, each cell at each stage $i = 0, 1, 2$ can at most go through one of the following events at an infinitesimally small time interval $\Delta t$:

1. Birth: Divide into two identical cells at a rate $\alpha_i$ with the probability $\alpha_i \Delta t + O(\Delta t^2)$,
2. Death: Die at a rate $\beta_i$ with the probability $\beta_i \Delta t + O(\Delta t^2)$,
3. Mutation: Mutate at a rate $\mu_i$ with the probability $\mu_i \Delta t + O(\Delta t^2)$,
4. Stay unchanged with the probability $1 - (\alpha_i + \beta_i + \mu_i)\Delta t + O(\Delta t^2)$.

Notice that for the normal stem cells, we do not consider birth and death rates, i.e., $\alpha_0 = \beta_0 = 0$, and for the malignant cells, we do not consider mutation ($\mu_3 = 0$), since we assume there are only three key mutations to malignancy as reported in [8]. Note, equations (1)–(4) are defined so that all short-term transition probabilities associated with events 1–4 can be expressed as their products. This attribute is necessary for deriving the corresponding system of ODEs in closed form. We will also neglect the higher order terms $O(\Delta t^2)$. We further assume that the cells are independent and their transition probability is independent of the time (i.e., they are time-homogeneous). Before proceeding with the rest of the derivation, we point out a useful equation called the Chapman-Kolmogorov equation. For a stochastic Markov process with the random variable $X(t)$ and an event $A$, we have:

$$Pr(X(t+s) \in A \mid X(0) = i) = \sum_{k=0}^{\infty} Pr(X(t+s) \in A \mid X(s) = k) * Pr(X(s) = k \mid X(0) = i). \tag{5}$$

So at $t + \Delta t$, we can carry out the following calculations:

$$\begin{aligned}
P_0(t+\Delta t) &= Pr(M(t+\Delta t) < M_{t+\Delta t} \mid I_0(0) = N_0, I_1(0) = 0, I_2(0) = 0, M(0) = 0) \\
&= \sum_{k_0,k_1,k_2,k_3} \Bigg[ Pr(M(t+\Delta t) < M_{t+\Delta t} \mid I_0(\Delta t) = N_0 + k_0, I_1(\Delta t) = k_1, I_2(\Delta t) = k_2, M(\Delta t) = k_3) \\
&\quad \times Pr(I_0(\Delta t) = N_0 + k_0, I_1(\Delta t) = k_1, I_2(\Delta t) = k_2, M(\Delta t) = k_3 \mid I_0(0) = N_0, I_1(0) = 0, I_2(0) = 0, M(0) = 0) \Bigg] \\
&= Pr(M(t+\Delta t) < M_{t+\Delta t} \mid I_0(\Delta t) = N_0, I_1(\Delta t) = 1, I_2(\Delta t) = 0, M(\Delta t) = 0)\mu_0 N_0 \Delta t \\
&+ Pr(M(t+\Delta t) < M_{t+\Delta t} \mid I_0(\Delta t) = N_0, I_1(\Delta t) = 0, I_2(\Delta t) = 0, M(\Delta t) = 0)(1 - \mu_0 N_0 \Delta t) \\
&= \mu_0 N_0 \Delta t P_0(t) P_1(t) + P_0(t) - \mu_0 N_0 \Delta t P_0(t).
\end{aligned}$$

The first equality comes from the Chapman-Kolmogorov equation. Notice that in the short time $\Delta t$, normal cells either mutate or stay unchanged. Hence, the infinite summation will only have two nonzero terms. Also, we assume that we start with $N_0$ normal cells, so their mutation rate will be $\mu_0 N_0$. The time homogeneity assumption allows us to write the probabilities in $[0, t]$ instead of $[\Delta t, t + \Delta t]$ (since they both have the same length), and the independence of normal cells from the first initiated cells leads to the decomposition $P_0(t)P_1(t)$. Now, all we need to do is subtract $P_0(t)$ from both sides, divide by $\Delta t$, and take the limit as $\Delta t \to 0$. We will get:

$$\frac{dP_0}{dt} = \mu_0 N_0 P_0(t)(P_1(t) - 1). \tag{6}$$

For $P_1(t + \Delta t)$ we have:

$$\begin{aligned}
P_1(t+\Delta t) &= Pr(M(t+\Delta t) < M_{t+\Delta t} \mid I_0(0) = N_0, I_1(0) = 1, I_2(0) = 0, M(0) = 0) \\
&= \sum_{k_0,k_1,k_2,k_3} \Bigg[ Pr(M(t+\Delta t) < M_{t+\Delta t} \mid I_0(\Delta t) = N_0 + k_0, I_1(\Delta t) = k_1, I_2(\Delta t) = k_2, M(\Delta t) = k_3) \\
&\quad \times Pr(I_0(\Delta t) = N_0 + k_0, I_1(\Delta t) = k_1, I_2(\Delta t) = k_2, M(\Delta t) = k_3 \mid I_0(0) = 0, I_1(0) = 1, I_2(0) = 0, M(0) = 0) \Bigg] \\
&= Pr(M(t+\Delta t) < M_{t+\Delta t} \mid I_0(\Delta t) = N_0, I_1(\Delta t) = 1, I_2(\Delta t) = 1, M(\Delta t) = 0)\mu_1 \Delta t \\
&+ Pr(M(t+\Delta t) < M_{t+\Delta t} \mid I_0(\Delta t) = N_0, I_1(\Delta t) = 2, I_2(\Delta t) = 0, M(\Delta t) = 0)\alpha_1 \Delta t \\
&+ Pr(M(t+\Delta t) < M_{t+\Delta t} \mid I_0(\Delta t) = N_0, I_1(\Delta t) = 0, I_2(\Delta t) = 0, M(\Delta t) = 0)\beta_1 \Delta t \\
&+ Pr(M(t+\Delta t) < M_{t+\Delta t} \mid I_0(\Delta t) = N_0, I_1(\Delta t) = 1, I_2(\Delta t) = 0, M(\Delta t) = 0)(1 - \mu_1 \Delta t - \alpha_1 \Delta t - \beta_1 \Delta t) \\
&= \mu_1 \Delta t P_1(t) P_2(t) + \alpha_1 \Delta t P_1(t)^2 + 1\beta_1 \Delta t + (1 - \mu_1 \Delta t - \alpha_1 \Delta t - \beta_1 \Delta t) P_1(t).
\end{aligned}$$

This time, we have four nonzero terms corresponding to FSMCs: birth, death, mutation, and no change. The probability multiplied by $\beta_1 \Delta t$ is equal to one because in case there are no cells in any compartments, the size of the tumor cells will always be zero, which is certainly smaller than a non-zero $M_{t+\Delta t}$. Once again, we subtract $P_1(t)$ from both sides, divide by $\Delta t$ and take the limit as $\Delta t \to 0$ to get:

$$\frac{dP_1}{dt} = \beta_1 - (\mu_1 + \alpha_1 + \beta_1)P_1(t) + \mu_1 P_1(t) P_2(t) + \alpha_1 P_1(t)^2. \tag{7}$$

The next equation is derived in exactly the same way.

$$\frac{dP_2}{dt} = \beta_2 - (\mu_2 + \alpha_2 + \beta_2)P_2(t) + \mu_2 P_2(t) P_3(t) + \alpha_2 P_2(t)^2. \tag{8}$$

To close the system, We either need an ODE for $P_3(t)$ or a function to replace it. As mentioned before, we assume that malignant cells go through a simple time-homogeneous birth-death process. We start by calculating the probability of having $n$ malignant cells at time $t$ (i.e., $p_n(t)$) given one malignant cell at time 0. It is easy to arrive at the following ODE using the same derivation logic as before for $p_n(t+\Delta t)$ and only considering birth and death transitions. This will lead to a recursive system of probabilities.

$$\begin{cases} \dfrac{dp_n}{dt} & = \alpha_3(n-1)p_{n-1} - (\alpha_3+\beta_3)np_n + \beta_3(n+1)p_{n+1} \qquad \text{for} \qquad n \geq 1, \\ \dfrac{dp_0}{dt} & = \beta_3 p_1. \end{cases} \tag{9}$$

To find $P_3(t)$ given in (4), we can solve (9) along with the initial conditions $p_i(0) = \delta_{i1}$, where $\delta$ is the Kronecker delta function. Thereafter, we can calculate $P_3(t) = \sum_{i=0}^{M_t-1} p_i(t)$. However, for even moderate values of $n$, this task becomes quite cumbersome. Instead, we use the moment-generating partial differential equation (PDE) of the process. Per [9, 7], it can be shown that for the time-homogeneous birth and death transitions with $M(0) = 1$, the moment-generating PDE is given by the following:

$$\frac{\partial \mathcal{M}}{\partial t} = (\alpha_3(e^\theta - 1) + \beta_3(e^{-\theta} - 1))\frac{\partial \mathcal{M}}{\partial \theta}, \tag{10}$$

with $\mathcal{M}(\theta, 0) = e^\theta$. Applying the separation of variables technique for PDEs, we can arrive at a solution for (10):

$$\mathcal{M}(\theta, t) = \frac{\alpha_3 e^{\beta_3 t+\theta} - \beta_3\left(-e^{\alpha_3 t} + e^{\beta_3 t} + e^{\alpha_3 t+\theta}\right)}{-\beta_3 e^{\beta_3 t} + \alpha_3\left(e^{\alpha_3 t} - e^{\alpha_3 t+\theta} + e^{\beta_3 t+\theta}\right)}. \tag{11}$$

This expression holds when $\alpha_3 \neq \beta_3$, which is what we assume in this paper. More precisely, we assume that malignant cells do not regress, i.e., $\alpha_3 > \beta_3$. We can get the probability-generating function by changing the variable $\theta = \ln x$.

$$\mathcal{P}(x, t) = \frac{\beta_3 + \beta_3 e^{(\alpha_3-\beta_3)t}(x-1) - \alpha_3 x}{\beta_3 + \alpha_3 e^{(\alpha_3-\beta_3)t}(x-1) - \alpha_3 x}, \tag{12}$$

Now that we have the probability-generating function, finding $p_n(t)$ is straightforward.

$$p_n(t) = \frac{1}{n!}\frac{\partial^n \mathcal{P}}{\partial x^n}\bigg|_{x=0} = (\alpha_3-\beta_3)^2 e^{(\alpha_3-\beta_3)t}\frac{(\alpha_3 e^{(\alpha_3-\beta_3)t} - \alpha_3)^{n-1}}{(\alpha_3 e^{(\alpha_3-\beta_3)t} - \beta_3)^{n+1}}. \tag{13}$$

Finally, we can calculate an expression for $P_3(t)$.

$$\begin{aligned} P_3(t) = \sum_{i=0}^{M_t-1} p_i(t) = 1 - \sum_{i=M_t}^{\infty} p_i(t) &= 1 - (\alpha_3-\beta_3)^2 e^{(\alpha_3-\beta_3)t}\sum_{i=M_t}^{\infty}\frac{(\alpha_3 e^{(\alpha_3-\beta_3)t} - \alpha_3)^{i-1}}{(\alpha_3 e^{(\alpha_3-\beta_3)t} - \beta_3)^{i+1}} \\ &= 1 - \frac{(\alpha_3-\beta_3)^2 e^{(\alpha_3-\beta_3)t}}{(\alpha_3 e^{(\alpha_3-\beta_3)t} - \beta_3)^2}\sum_{i=M_t}^{\infty}\left(\frac{\alpha_3 e^{(\alpha_3-\beta_3)t} - \alpha_3}{\alpha_3 e^{(\alpha_3-\beta_3)t} - \beta_3}\right)^{i-1}. \end{aligned}$$

Notice that the fraction inside the infinite sum is always less than one if $\alpha_3 > \beta_3$. So, by way of geometric series, we get:

$$P_3(t) = 1 - \frac{(\alpha_3-\beta_3)\alpha_3^{M_t-1}\{1 - e^{(\beta_3-\alpha_3)t}\}^{M_t-1}}{\{\alpha_3 - \beta_3 e^{(\beta_3-\alpha_3)t}\}^{M_t}}. \tag{14}$$

So far, we have the following ODE system given by equations (6)-(8):

$$\begin{aligned} \frac{dP_0}{dt} &= \mu_0 N_0 P_0(t)(P_1(t) - 1), \\ \frac{dP_1}{dt} &= \beta_1 - (\mu_1 + \alpha_1 + \beta_1)P_1(t) + \mu_1 P_1(t)P_2(t) + \alpha_1 P_1(t)^2, \\ \frac{dP_2}{dt} &= \beta_2 - (\mu_2 + \alpha_2 + \beta_2)P_2(t) + \mu_2 P_2(t)P_3(t) + \alpha_2 P_2(t)^2. \end{aligned}$$

Where $P_3(t)$ is given by (14) and the initial conditions, based on the definition of each probability, are $\{P_0(0) = 1, P_1(0) = 1, P_2(0) = 1\}$. Now, to solve for the instantaneous hazard numerically and simultaneously, we adopt the following change of variables.

$$\left\{x_1(t) = P_0(t)\ ,\ x_2(t) = -\frac{d}{dt}\ln(x_1(t))\ ,\ x_3(t) = P_1(t)\ ,\ x_4(t) = x_3'(t)\ ,\ x_5(t) = P_2(t)\ ,\ x_6(t) = x_5'(t)\ ,\ f(t) = P_3(t)\right\}.$$

With these, we get the system of equations given in the main text with initial conditions $[x_1, x_2, x_3, x_4, x_5, x_6] = [1, 0, 1, 0, 1, 0]$.

$$\frac{dx_1}{dt} = \mu_0 N_0 x_1(x_3 - 1), \tag{15}$$

$$\frac{dx_2}{dt} = -\mu_0 N_0 x_4, \tag{16}$$

$$\frac{dx_3}{dt} = \beta_1 - (\alpha_1 + \beta_1 + \mu_1)x_3 + \mu_1 x_3 x_5 + \alpha_1 x_3^2, \tag{17}$$

$$\frac{dx_4}{dt} = -(\alpha_1 + \beta_1 + \mu_1)x_4 + \mu_1 x_4 x_5 + \mu_1 x_3 x_6 + 2\alpha_1 x_3 x_4, \tag{18}$$

$$\frac{dx_5}{dt} = \beta_2 - (\alpha_2 + \beta_2 + \mu_2)x_5 + \mu_2 f(t) x_5 + \alpha_2 x_5^2, \tag{19}$$

$$\frac{dx_6}{dt} = -(\alpha_2 + \beta_2 + \mu_2)x_6 + \mu_2 f'(t) x_5 + \mu_2 f(t) x_6 + 2\alpha_2 x_6 x_5. \tag{20}$$

For the classic model, we set $\alpha_3 = \beta_3 = 0$, and for the simple birth process, we let $\beta_3 = 0$ in $f(t)$. This concludes our derivation.

### Structural identifiability

The classic model that considers the hazard as the occurrence of the first malignancy is not structurally identifiable [10]. Given the relationship between survival and hazard, the information they require is inherently the same. So, for simplicity, we investigate (6)-(8) instead of (15)-(20). We follow the same procedure as Brouwer et al. [10] by constructing a monic algebraic equation with respect to the input and output and their derivatives in decreasing order, also known as an input-output equation. We start by solving (6) for $P_1(t)$ with respect to $P_0(t)$ and its derivative. We then take the derivative of the resulting expression to get $P_1'(t)$. Substituting these in (7), we can find an expression for $P_2(t)$ with respect to $P_0(t)$ and its derivatives. Plugging all of that into (8) will give us an algebraic equation with respect to $P_0(t)$ and its derivatives and $P_3(t)$. Table S2 shows the terms in decreasing order with their coefficients.

If we assume $P_3(t)$ is a known input (i.e., $\beta_3 = 0$ and $\alpha_3$ is a known value from the literature), then the algebraic equation we discussed will be ordered according to Table S2. The common way of ordering is $P_0^{(3)} > P_0'' > P_0' > P_0 > P_3$. So, the highest order belongs to the term $P_0^2 P_0' P_0^{(3)}$. This term has to have a coefficient of 1 before we can consider this as an input-output equation. This is how the parameters are identified structurally:

- $\mu_0$: immediately identifiable as the sole coefficient of the term with order 2.
- $\mu_2$: immediately identifiable as the sole coefficient of the term with order 5.
- $\mu_1$: Since $\mu_0$ and $\mu_2$ are identifiable then $\mu_1$ can be identified according to the term with order 16.
- $\alpha_1$: Since $\mu_0$ and $\mu_2$ are identifiable, then $\alpha_1$ can be identified according to the term with order 10.
- $\alpha_2$: Since $\mu_0$, $\mu_1$ and $\alpha_1$ are identifiable, then $\alpha_2$ can be identified according to the term with order 9.
- $\beta_1$: Since $\mu_0$, $\mu_1$, $\mu_2$, and $\alpha_1$ are identifiable, then $\beta_1$ can be identified according to the term with order 14.
- $\beta_2$: everything else has been identified so $\beta_2$ can be identified via the term with order 8.

The assumption that $P_3$ is a known input immediately identified $\mu_1$ and $\mu_2$ independently. Also, it directly helped with the identifiability of $\alpha_1$ and $\beta_1$. In the case of the classic model where $\alpha_3 = \beta_3 = 0$ (i.e., $P_3(t) = 0$), terms with orders 5, 7, 10, 12, 14, and 16 vanish. Then $\mu_1$, $\mu_2$ and $\alpha_1$, $\beta_1$, $\alpha_2$ and $\beta_2$ will only appear in combinations and not independently, see conjecture 1 in [10]. Moreover, if we consider nonzero and unknown parameters $\alpha_3$ and $\beta_3$, then $P_3$ will no longer play a role in the "terms" column of Table S2. For example, the table's terms of order 5 and 6 are only separated due to the existence of $P_3$ in the order 5 term. If $P_3$ is not a known input, it will multiply by the coefficient of order 5 and add to the coefficients of order 6. This is true for terms with orders 10 and 11, 14 and 15, and 16 and 17. Since $P_3$ is a highly nonlinear function of $\alpha_3$ and $\beta_3$ this will only increase the non-identifiability of all the parameters. For these reasons, we use Equation 11 in the main text to ensure that the whole system is identifiable.

Table S2: Terms and coefficients of the input-output equation of (6)-(8). The superscripts in parentheses denote derivatives.

| Order | Terms | Coefficients |
|---|---|---|
| 1 | $P_0^2 P_0' P_0^{(3)}$ | 1 |
| 2 | $P_0^3 P_0^{(3)}$ | $\mu_0$ |
| 3 | $P_0^2 (P_0'')^2$ | $-1 - \frac{\alpha_2}{\mu_1}$ |
| 4 | $P_0 (P_0')^2 P_0''$ | $-1 - \frac{2\alpha_2}{\mu_1} + \frac{2\alpha_1\alpha_2}{\mu_1\mu_0}$ |
| 5 | $P_3 P_0^2 P_0' P_0''$ | $-\mu_2$ |
| 6 | $P_0^2 P_0' P_0''$ | $-3\mu_0 + \mu_2 + \beta_2 - \alpha_2 - 2\alpha_1 + \frac{2\alpha_1\alpha_2}{\mu_1} - \frac{2\alpha_2\beta_1}{\mu_1}$ |
| 7 | $P_3 P_0^3 P_0''$ | $-\mu_2\mu_0$ |
| 8 | $P_0^3 P_0''$ | $\mu_2\mu_0 + \beta_2\mu_0 + \beta_1\mu_0 - \alpha_2\mu_0 - \alpha_1\mu_0$ |
| 9 | $(P_0')^4$ | $1 - \frac{\alpha_2}{\mu_1} + \frac{\alpha_1}{\mu_0} + \frac{2\alpha_1\alpha_2}{\mu_0\mu_1} - \frac{\alpha_1^2\alpha_2}{\mu_0^2\mu_1}$ |
| 10 | $P_3 P_0 (P_0')^3$ | $\mu_2 + \frac{\alpha_1\mu_2}{\mu_0}$ |
| 11 | $P_0 (P_0')^3$ | $2\mu_0 - \mu_2 - \beta_2 + \alpha_2 + 2\alpha_1 + \frac{2\alpha_2\beta_1}{\mu_1} - \frac{2\alpha_1\alpha_2}{\mu_1} - \frac{\alpha_1\mu_2}{\mu_0^2} - \frac{\alpha_1\beta_2}{\mu_0} + \frac{\alpha_1\alpha_2}{\mu_0} + \frac{2\alpha_1\alpha_2\beta_1}{\mu_1\mu_0} - \frac{2\alpha_1^2\alpha_2}{\mu_1\mu_0}$ |
| 12 | $P_3 P_0^2 (P_0')^2$ | $\mu_0\mu_2 - \mu_1\mu_2 - \beta_1\mu_2 + 2\alpha_1\mu_2$ |
| 13 | $P_0^2 (P_0')^2$ | $-\mu_0\mu_2 - \beta_2\mu_0 - \beta_1\mu_0 + \alpha_2\mu_0 + \alpha_1\mu_0 + \mu_1\mu_2 + \beta_1\mu_2 - 2\alpha_1\mu_2 + \beta_1\beta_2 - 2\alpha_1\beta_2 - \alpha_2\beta_1 + 2\alpha_1\alpha_2 - \frac{\alpha_2\beta_1^2}{\mu_1} + \frac{2\alpha_1\alpha_2\beta_1}{\mu_1} - \frac{\alpha_1^2\alpha_2}{\mu_1}$ |
| 14 | $P_3 P_0^3 P_0'$ | $-2\mu_1\mu_2\mu_0 - \beta_1\mu_2\mu_0 + \alpha_1\mu_2\mu_0$ |
| 15 | $P_0^3 P_0'$ | $2\mu_1\mu_2\mu_0 + \beta_1\mu_2\mu_0 - \alpha_1\mu_2\mu_0 + \beta_1\beta_2\mu_0 - \alpha_1\beta_2\mu_0 - \alpha_2\beta_1\mu_0 + \alpha_1\alpha_2\mu_0$ |
| 16 | $P_3 P_0^4$ | $-\mu_1\mu_2\mu_0^2$ |
| 17 | $P_0^4$ | $\mu_1\mu_2\mu_0^2$ |

### Age at menarche

Table S3: Age at menarche for different cohorts extracted from NHANES

| Cohort | Age at menarche |
|---|---|
| 1930-1934 | 12.92 (CI: 12.8-13.03) |
| 1935-1939 | 12.97 (CI: 12.87-13.08) |
| 1940-1944 | 12.75 (CI: 12.63-12.88) |
| 1945-1949 | 12.67 (CI: 12.56-12.78) |
| 1950-1954 | 12.69 (CI: 12.57-12.80) |
| 1955-1959 | 12.72 (CI: 12.61-12.83) |
| 1960-1964 | 12.86 (CI: 12.75-12.97) |
| 1965-1969 | 12.76 (CI: 12.66-12.87) |
| 1970-1974 | 12.62 (CI: 12.52-12.72) |
| 1975-1979 | 12.51 (CI: 12.41-12.60) |
| 1980-1984 | 12.52 (CI: 12.43-12.61) |
| 1985-1989 | 12.49 (CI: 12.4-12.57) |
| 1990-1994 | 12.35 (CI: 12.24-12.45) |

### Sensitivity Analysis

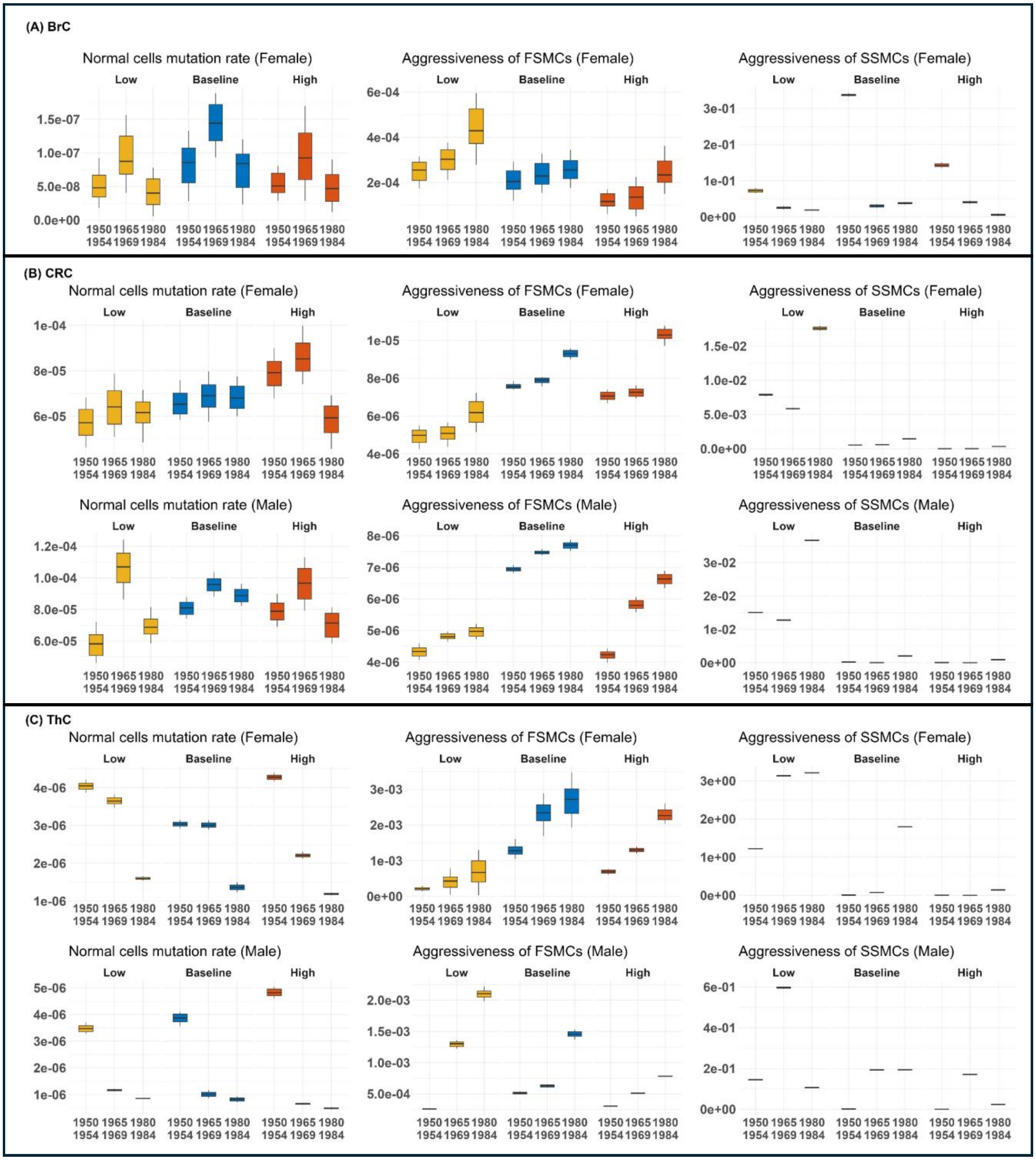


Figure S2: Sensitivity of parameter estimation to variations in $\alpha_3$. The distribution of estimated parameters is shown for three values of $\alpha_3$: Low ($\alpha_3$ reduced by 25% relative to the baseline), Baseline (values obtained from literature), and High ($\alpha_3$ increased by 25% relative to the baseline). Results are presented for (A) BrC, (B) CRC, and (C) ThC, and three cohorts of 1950-1954, 1965-1969, and 1980-1984. The parameters shown here are i) Normal cell mutation rate $\mu_0$; ii) Aggressiveness of the first-stage mutated cells (FSMCs) $\mu_1 \times (\alpha_1 - \beta_1)$; and iii) Aggressiveness of the second-stage mutated cells (SSMCs) $\mu_2 \times (\alpha_2 - \beta_2)$.

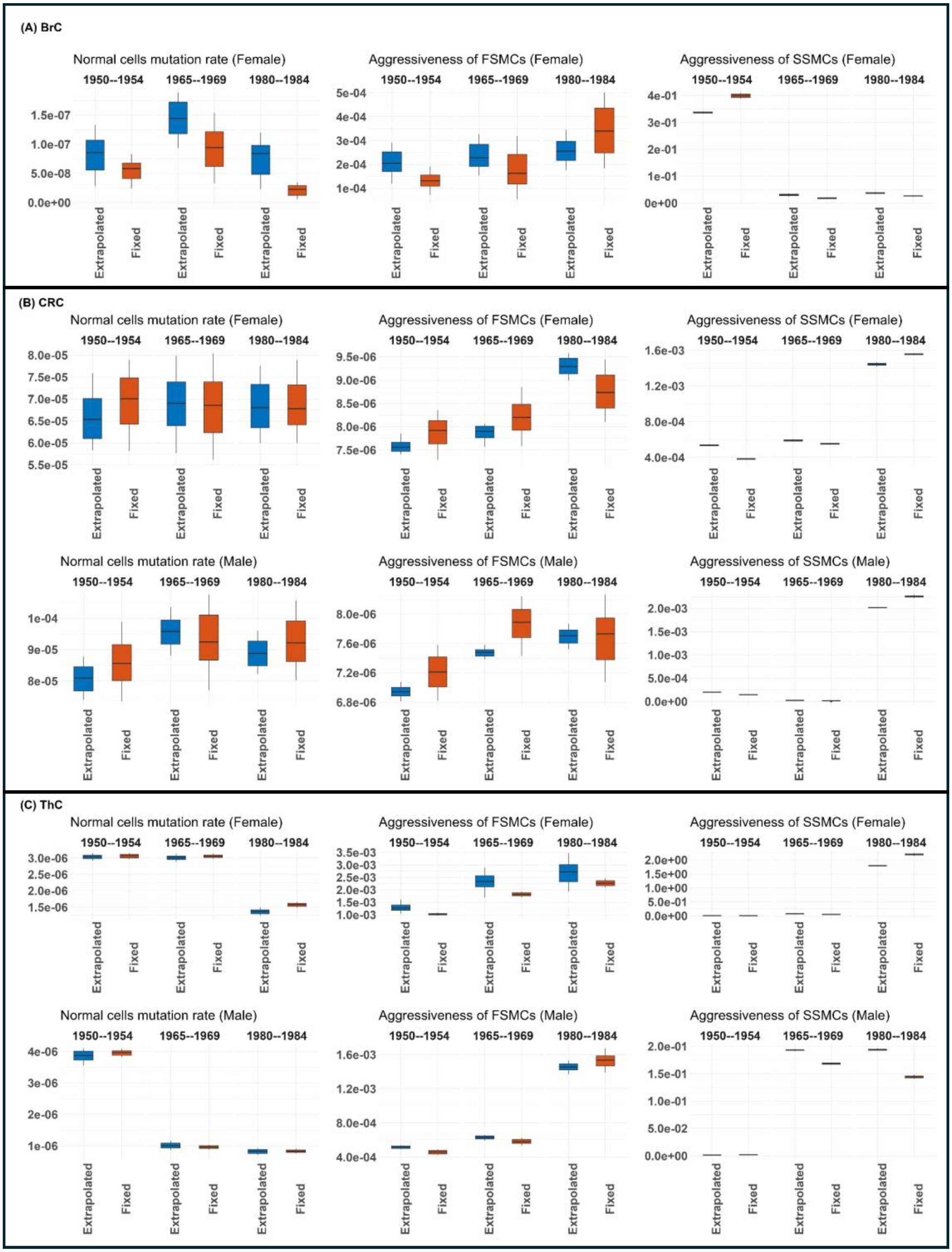


Figure S3: Comparison between estimated parameters for two cases of tumor-size-at-diagnosis input. The "Extrapolated" case is similar to the results in Figure 4 of the main manuscript, where missing tumor-size-at-diagnosis data are filled in via extrapolation. For the "Fixed" case, we filled in the missing data with a fixed value (i.e., the earliest available data).

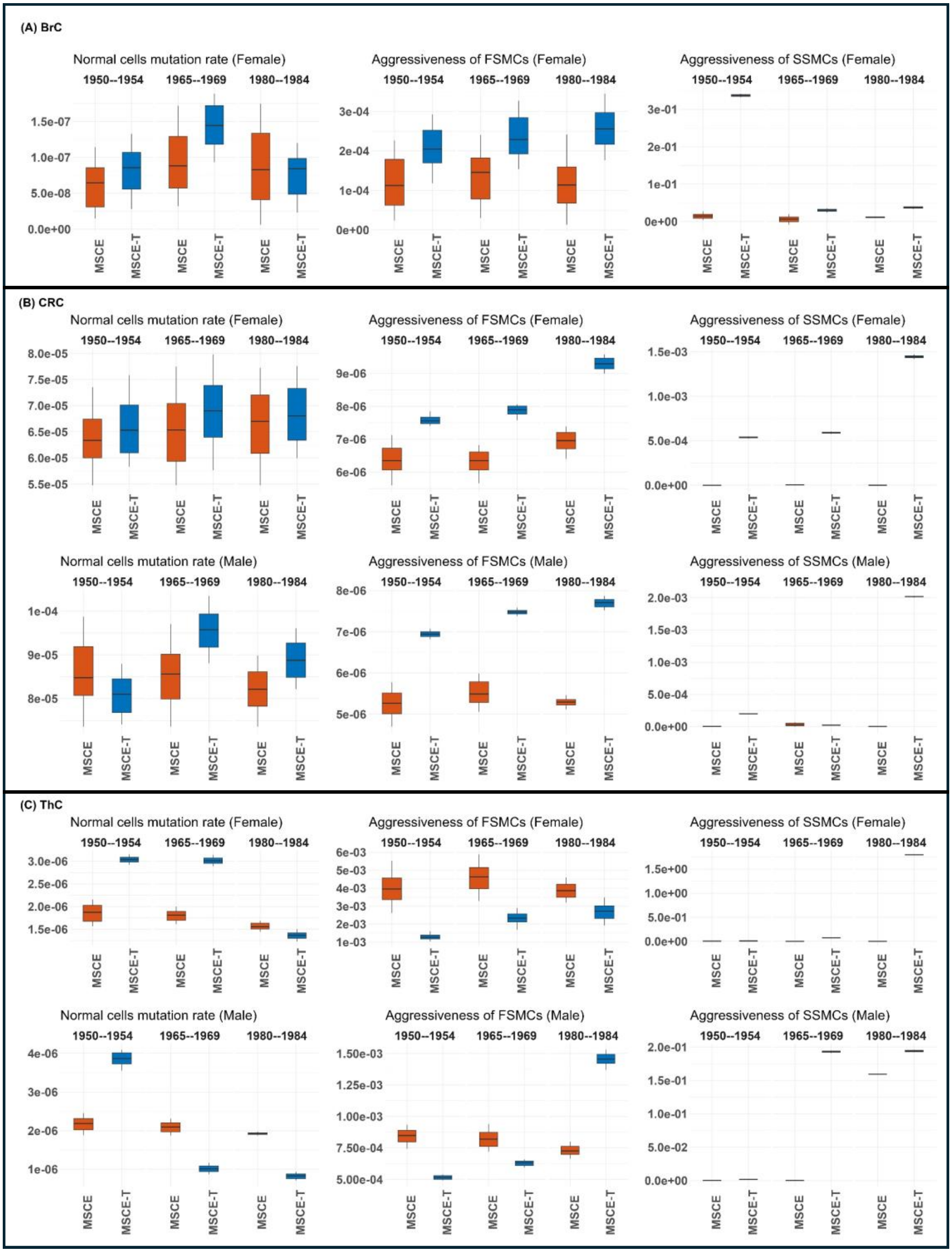


Figure S4: Comparison between three estimated parameter distributions corresponding to the MSCE model (orange bars) and the MSCE-T model (blue bars) for three types of cancer: (A) BrC, (B) CRC, and (C) ThC. The cohorts chosen for this comparison are 1950-1954, 1965-1969, and 1980-1984. The parameters shown here are i) Normal cell mutation rate $\mu_0$; ii) Aggressiveness of the first-stage mutated cells (FSMCs) $\mu_1 \times (\alpha_1 - \beta_1)$; and iii) Aggressiveness of the second-stage mutated cells (SSMCs) $\mu_2 \times (\alpha_2 - \beta_2)$.

Table S4: Comparison between the Negative Log Likelihood (NLL) and Akaike Information Criterion (AIC) values associated with Figure S4.

| Cancer Type | Sex | Cohort (Years) | MSCE-T | MSCE |
|---|---|---|---|---|
| BrC | Female | 1950-1954 | NLL: 52.4 & AIC: 118.7 | NLL: 54.1 & AIC: 122.2 |
| | | 1965-1969 | NLL: 58.3 & AIC: 130.6 | NLL: 60.9 & AIC: 135.8 |
| | | 1980-1984 | NLL: 47.1 & AIC: 108.2 | NLL: 47.6 & AIC: 109.2 |
| CRC | Female | 1950-1954 | NLL: 55.1 & AIC: 124.2 | NLL: 62.1 & AIC: 138.2 |
| | | 1965-1969 | NLL: 72.6 & AIC: 159.2 | NLL: 81.1 & AIC: 176.2 |
| | | 1980-1984 | NLL: 26.5 & AIC: 67.0 | NLL: 27.1 & AIC: 68.2 |
| CRC | Male | 1950-1954 | NLL: 57.0 & AIC: 128.0 | NLL: 64.2 & AIC: 142.4 |
| | | 1965-1969 | NLL: 64.9 & AIC: 143.8 | NLL: 73.5 & AIC: 161.0 |
| | | 1980-1984 | NLL: 26.4 & AIC: 66.8 | NLL: 28.8 & AIC: 71.6 |
| ThC | Female | 1950-1954 | NLL: 89.4 & AIC: 192.8 | NLL: 143.1 & AIC: 300.2 |
| | | 1965-1969 | NLL: 276.9 & AIC: 567.8 | NLL: 298.2 & AIC: 610.4 |
| | | 1980-1984 | NLL: 174.1 & AIC: 362.2 | NLL: 178.0 & AIC: 370.0 |
| ThC | Male | 1950-1954 | NLL: 56.5 & AIC: 127.0 | NLL: 72.4 & AIC: 158.8 |
| | | 1965-1969 | NLL: 128.6 & AIC: 271.2 | NLL: 146.3 & AIC: 306.6 |
| | | 1980-1984 | NLL: 69.7 & AIC: 153.4 | NLL: 73.6 & AIC: 161.2 |

### Anaplastic carcinoma vs papillary adenocarcinoma

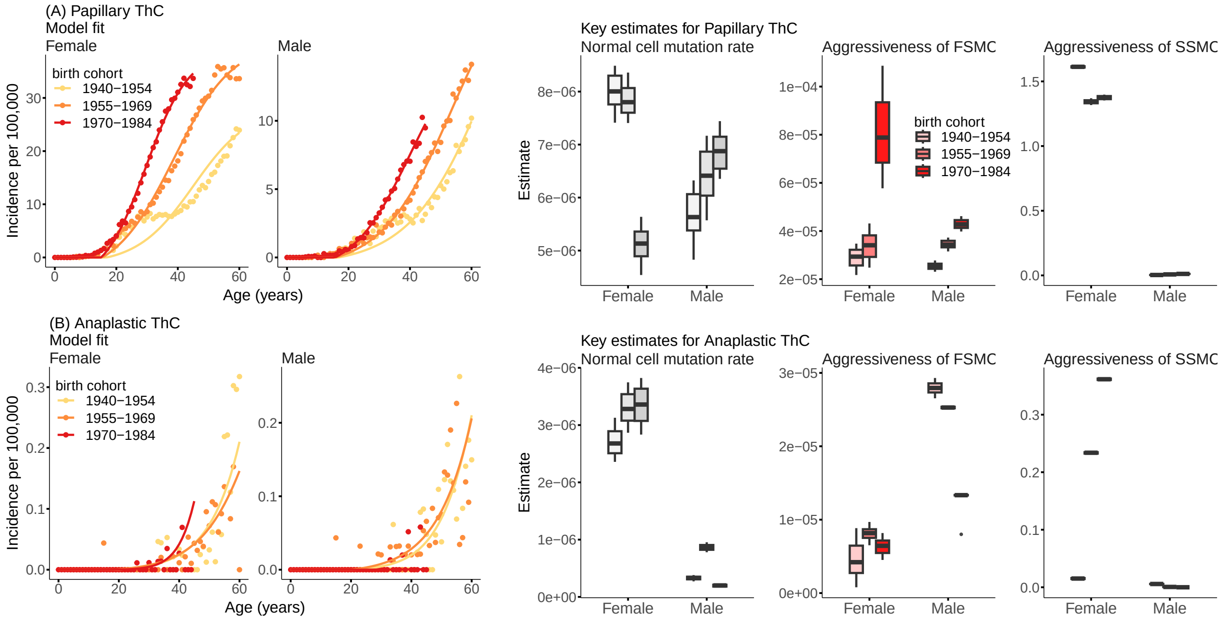


Figure S5: Comparison between the model fit and distribution of parameters estimated for two different subtypes of thyroid cancer (ThC): (A) Papillary adenocarcinoma and (B) Anaplastic carcinoma. The parameters shown here are i) Normal cell mutation rate $\mu_0$; ii) Aggressiveness of the first-stage mutated cells (FSMCs) $\mu_1 \times (\alpha_1 - \beta_1)$; and iii) Aggressiveness of the second-stage mutated cells (SSMCs) $\mu_2 \times (\alpha_2 - \beta_2)$. Because the incidence is very low for anaplastic ThC, for this analysis, we aggregate the data for 15-year cohorts (i.e.,1940-1954, 1955-1969, and 1970-1984) to reduce observational noise.

### Sojourn Times

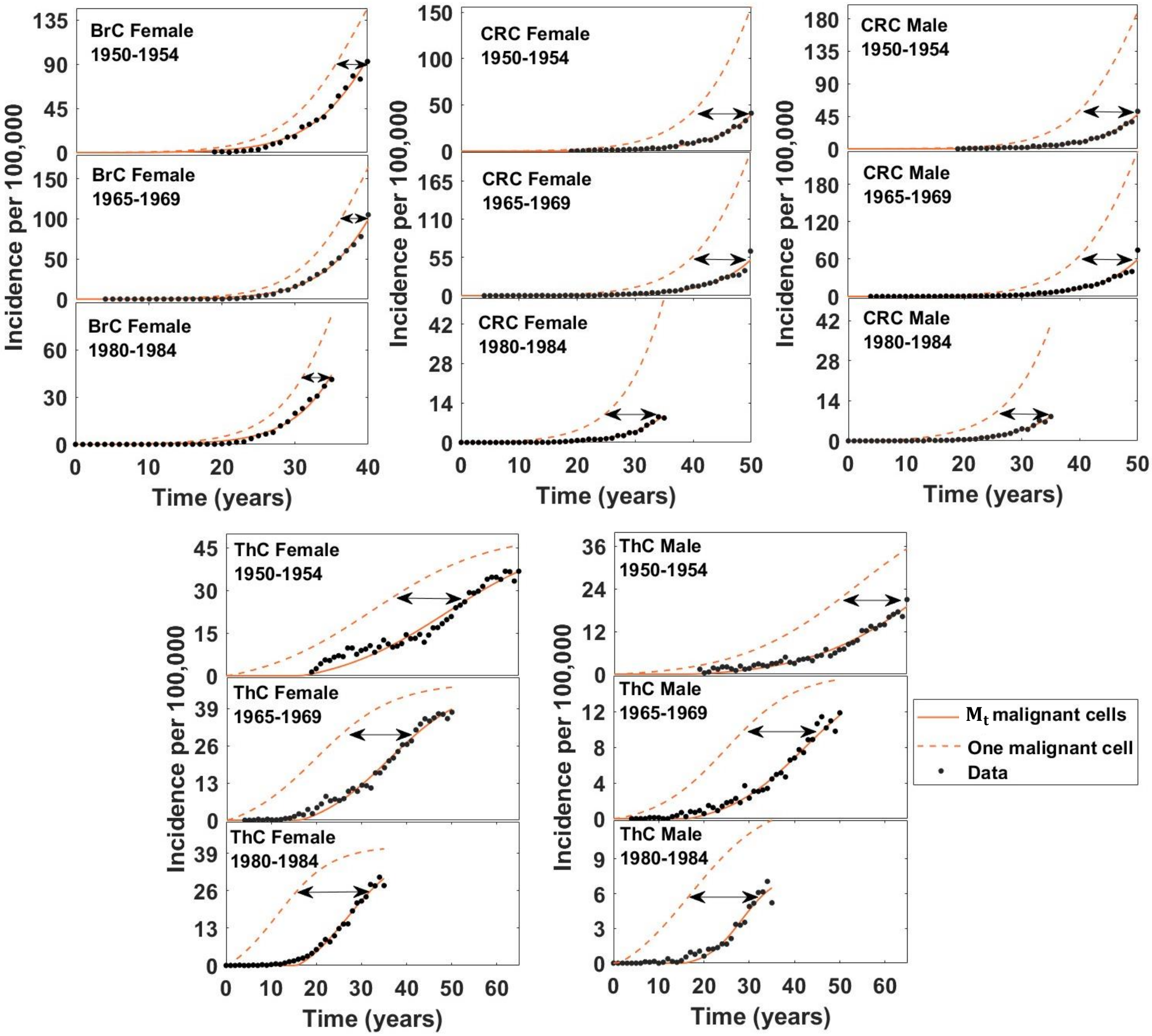


Figure S6: Illustration of sojourn time estimation. Sojourn time is the interval between the occurrence of a single malignant cell and when a clinically detectable number of malignant cells is present (i.e., $M_t$). The arrows show an instance of this time interval for a given level of incidence. The mean sojourn time is determined by averaging the sojourn times, restricted to cases for which relevant clinical data are available.
